# Distinguishing evidence of absence from absence of evidence in claims of safety: A methodological study with application to diagnostic hysteroscopy in endometrial cancer

**DOI:** 10.64898/2026.11.20.26368573

**Authors:** Rodrigo Alliende-Ferrada, Belén Mena, Andrés Ruz, Romina Benavides, Elisa Larraín, Ignacia Inostroza, Pablo Torres-Chomon

**Affiliations:** Gynecologic Oncology Unit, Hospital Padre Hurtado, Santiago, Chile; Universidad de Chile, Santiago, Chile; Faculty of Engineering, Pontificia Universidad Católica de Chile, Santiago, Chile; Universidad del Desarrollo, Santiago, Chile (medical student); Universidad Europea de Valencia, Valencia, Spain (MSc Public Policy); Unidad de Ginecología Oncológica, Hospital Barros Luco Trudeau, Santiago, Chile

## Abstract

**Background:** Claims that an intervention is safe often rest on a nonsignificant difference, confusing absence of evidence with evidence of absence; an integrated, reproducible procedure for auditing them is lacking. We developed and evaluated one, applying it to diagnostic hysteroscopy in endometrial cancer, which published syntheses describe as oncologically safe.

**Methods and findings:** The framework’s eight steps verify each number, test the direction of confounding, require time-to-event analysis, bound detectability and apply one information-adjusted boundary to claims of harm and of safety (verdicts: harm established, safety established, inconclusive). The rule was fixed before a retrospective test on minimally invasive radical hysterectomy for cervical cancer, later assessed in a randomized trial, and evaluated analytically and by simulation. We audited 18 comparative hysteroscopy studies: 12 with peritoneal cytology (4,078 patients), 5 with time-to-event recurrence. With a true hazard ratio (HR) of 1.25 and the information available for hysteroscopy, inferring safety from nonsignificance gave false reassurance in 34% of analyses and the rule in 1%; with adequate information and no harm, the rule established safety in 80%. Pre-trial evidence on minimally invasive radical hysterectomy proved inconclusive, not safe. For hysteroscopy, controlling for disease extent revealed an association with positive cytology (odds ratio 2.86, 95% confidence interval [CI] 1.24–6.62) absent from crude comparisons; recurrence data (HR 1.17, Hartung–Knapp 95% CI 0.90–1.52) held 71% of required information and were inconclusive at margins of 1.10–1.40; and cytology-mediated harm would be undetectable in low-risk disease. Limitations include development on the case to which it is applied, a single author-selected retrospective test, no independent or prospective validation, and reliance on published observational data.

**Conclusions:** The evidence neither establishes nor excludes oncologic harm from diagnostic hysteroscopy; the proposed precautions also involve clinical judgment. Claims of safety should state the harm they exclude; the framework, provisional pending independent validation, makes that check explicit and reproducible.

**Author summary:** *Why was this study done?:*

- Claims that a test or operation is safe often rest on studies that found no statistically significant difference, which does not show that a clinically important harm has been excluded.
- Diagnostic hysteroscopy is widely used to investigate abnormal uterine bleeding and is described as oncologically safe in endometrial cancer, although the fluid that distends the uterus can carry tumor cells into the abdomen.
- Tools exist for parts of this problem, such as rating the certainty of evidence, but there has been no single, reproducible procedure for deciding whether a claim of safety is supported.

*What did the researchers do and find?:*

- We developed an eight-step framework that traces every number to its source, tests the direction of bias, asks whether the harm could have been detected at all, and applies one rule, adjusted for the amount of evidence, to claims of harm and of safety alike.
- When a clinically important harm existed, inferring safety from a nonsignificant result gave false reassurance in 34% to 91% of analyses based on limited evidence, against 1% or less with the framework, which also classified as unproven a pre-trial claim of safety for minimally invasive radical hysterectomy that a randomized trial later contradicted.
- In 18 studies of hysteroscopy, positive peritoneal cytology was more frequent once the extent of disease was taken into account, and recurrence data, with 71% of the information needed, could neither establish nor exclude a clinically important increase.

*What do these findings mean?:*

- A claim of safety should state the harm it rules out: with the two limits of a published 95% confidence interval, one table gives the verdict and its meaning within minutes, and a worksheet, a one-page committee summary, a free online calculator and open code support a full audit by clinicians, reviewers and guideline panels.
- For hysteroscopy, harm is neither shown nor excluded; because such harm is plausible and serious, the findings support precautions rather than a conclusion of harm, although these precautions, unlike the verdict, also rest on clinical judgment: the lowest effective distension pressure in every case, and curettage first when cancer is likely and preoperative information is incomplete, scaled to an expected 1.4 to 31 additional positive cytology results per 1,000 procedures and to be revised as evidence accumulates.
- The framework was developed on the case to which it is applied and tested retrospectively on one further case, so it should be regarded as provisional until others apply it; a protocol, a reporting list and a minimal check for journals and guideline panels are provided for them.

## Introduction

Claims that an intervention is safe are among the most consequential statements in clinical evidence, because they remove an option from scrutiny. Such claims are often based on the absence of a statistically significant difference, yet a nonsignificant result is not evidence of no difference unless the confidence interval excludes clinically important effects and the evidence was large enough to detect them [1]. Misinterpretation of nonsignificant results remains common even among randomized trials published in high-impact journals [2], and it persists in current syntheses. A 2022 meta-analysis of uterine manipulators in minimally invasive surgery for endometrial cancer found no significant difference in recurrence (risk ratio 1.11, 95% confidence interval [CI] 0.71–1.74), concluded that manipulators do not increase recurrence and questioned whether further studies were needed [3]; yet that interval was compatible with a 74% increase in risk, and a 2026 meta-analysis of time-to-event data found shorter disease-free survival with manipulator use (hazard ratio 1.18, 95% CI 1.01–1.38) [4]. In observational evidence about harm, two further problems compound the error. Confounding by indication can bias comparisons toward no effect [5], and numbers transcribed between secondary sources can drift from what primary studies reported [6,7]. Because systematic reviews and meta-analyses are numerous and often overlap [8], an inference that has not been checked against primary data can be carried forward into later syntheses and guidance.

Existing methods address parts of this problem. The Grading of Recommendations Assessment, Development and Evaluation (GRADE) approach rates the certainty of evidence and treats confidence intervals that include appreciable harm as imprecise [9,10]; the Risk Of Bias In Non-randomised Studies of Interventions (ROBINS-I) tool assesses bias in individual nonrandomized studies [11]; trial sequential analysis relates the information accrued to the information required [12]; procedures for statistical and clinical significance combine several of these elements for meta-analyses of randomized trials [13]; equivalence testing formalizes how the absence of an effect can be supported [14]; and E-values quantify the confounding needed to explain an association away [15]. What is missing is an integrated, reproducible procedure for one specific and common question: does the evidence establish the absence of clinically important harm, or merely fail to show harm? Such a procedure must verify the numbers before analyzing them, predict and test the direction of bias, ask whether the harm could have been detected at all, and connect the result to a decision rule that neither dismisses uncertainty nor exaggerates it.

That decision rule is the domain of the precautionary principle. In a widely cited formulation, precaution involves preventive action in the face of uncertainty, a shift of the burden of proof to the proponents of an activity, exploration of alternatives and broader participation in decisions [16]. Critics have argued that strong versions of the principle are incoherent, because inaction and alternatives also carry risks [17], and others have pointed out that precaution applies to precautionary measures themselves [18]. Steel’s account answers these objections by requiring a credible scientific case for harm (knowledge condition), a threat of sufficient seriousness (harm condition) and a proportionate response; a measure is proportionate when it is not itself ruled out by the same principle (consistency) and is the least burdensome among those that are not (efficiency) [19]. These elements can be made operational, but they are rarely applied in clinical evidence synthesis.

Diagnostic hysteroscopy in endometrial cancer is a suitable test case. Joint statements of gynecologic societies favor visually directed over blind endometrial sampling wherever feasible [20,21], and cancer is present in about 9% of women with postmenopausal bleeding [22] and in 13.1% of hysteroscopic polypectomies in women aged 70 years or older who bleed [23].

Distension can carry fluid through the fallopian tubes above a threshold pressure [24], and transtubal passage of viable tumor cells has been documented after hysteroscopy in women with endometrial cancer [25]. Meta-analyses have found more positive peritoneal cytology after hysteroscopy [26,27], and positive cytology is associated with worse outcomes, including in low-risk disease [28,29]. Yet published syntheses have concluded that hysteroscopy is oncologically safe, mainly because survival did not differ significantly with and without it [30–32].

We aimed to develop an explicit and reproducible framework for auditing claims of safety; to evaluate its central decision rule by exact calculation and simulation; to test it retrospectively, with the rule fixed in advance, on a question whose answer is now known; to apply it to diagnostic hysteroscopy in endometrial cancer; and to provide the data, code, a worksheet, a calculator that runs in any web browser, a table that applies the central step to any published confidence interval without software and a protocol for independent audits, so that clinicians, reviewers and guideline panels can apply it to other claims. The contribution is one of integration rather than of new statistics: a single decision rule, applied symmetrically to claims of harm and of safety against a margin set in advance, preceded by checks on the numbers, the direction of bias, the effect scale and detectability, and followed by a decision that states what evidence would change it. The rule itself is not new: it is the single-analysis boundary of trial sequential analysis, applied with equal stringency to claims of harm and of safety. What the framework adds is the order of the checks around it and a strict separation between the verdict, which the locked rule computes, and the precautions, which require clinical judgment and are reported with the assumptions on which they depend.

## Methods

### Study design and analysis plan

This is a methodological study with three components: development of an audit framework, evaluation of its central decision rule, and two applications, a retrospective test on a question whose answer is now known and an audit of diagnostic hysteroscopy in endometrial cancer. No prospective protocol was written or registered. The analyses were determined in the following sequence. First, the eight steps were developed iteratively while the hysteroscopy evidence was being re-analyzed. Second, before the retrospective test, the decision rule of Step 5 was specified, implemented in code and locked: the margin of 1.25, the information required for 80% power at a two-sided α of 0.05, the boundary function and the definitions of the three verdicts. Third, the locked rule was applied unchanged to the retrospective test and to the evaluation of its operating characteristics; nothing in the rule was modified after the retrospective results were seen. The rule contains no parameter estimated from data: its constants are conventional (the GRADE threshold for appreciable harm, 80% power, a two-sided α of 0.05 and an O’Brien–Fleming-type boundary), which leaves little room for tuning. Development and testing of the rule were therefore kept apart: the rule was fixed before it was applied to minimally invasive radical hysterectomy, the only empirical test of the rule reported here, whereas the hysteroscopy application illustrates the complete framework, supplied none of the rule’s constants and is not offered as a test of it.

We regard the locked Step 5 rule, the evaluation of its operating characteristics and the retrospective test as prespecified analyses. Four sets of analyses were defined after the main results had been seen and are reported as exploratory: (1) the grouping of cytology studies by whether disease extent was controlled, which is based on a design feature of each comparison but was defined during the analysis; (2) sensitivity analyses for the margin, for the prognostic weight of induced positive cytology and for the pretest probability of cancer; (3) boundaries calibrated recursively for repeated updates of the same evidence; and (4) the presentation of Step 5 as an audit interval and as a table of confidence limits, both algebraically equivalent to the locked rule. None of these exploratory analyses changed a Step 5 verdict reported for either application.

The hysteroscopy application is an illustrative audit, not a systematic review, and it was not registered. Its structured search and selection are reported according to the Preferred Reporting Items for Systematic Reviews and Meta-Analyses (PRISMA) 2020 statement [33] (S1 Checklist); the checklist does not apply to the framework itself. Only published aggregate data were used, so ethics approval and informed consent were not required.

### The audit framework

The framework applies to a claim that an intervention does not cause clinically important harm. Its eight steps answer three questions in turn (Fig 1; Table 1): is the evidence what it is said to be (Steps 1 and 2), does it support the claim (Steps 3 to 6), and what follows for practice (Steps 7 and 8). Each step has a method, an output and, where possible, an explicit decision rule. Formulas are given in S1 Appendix and a blank worksheet in S1 File.

**Fig 1.**
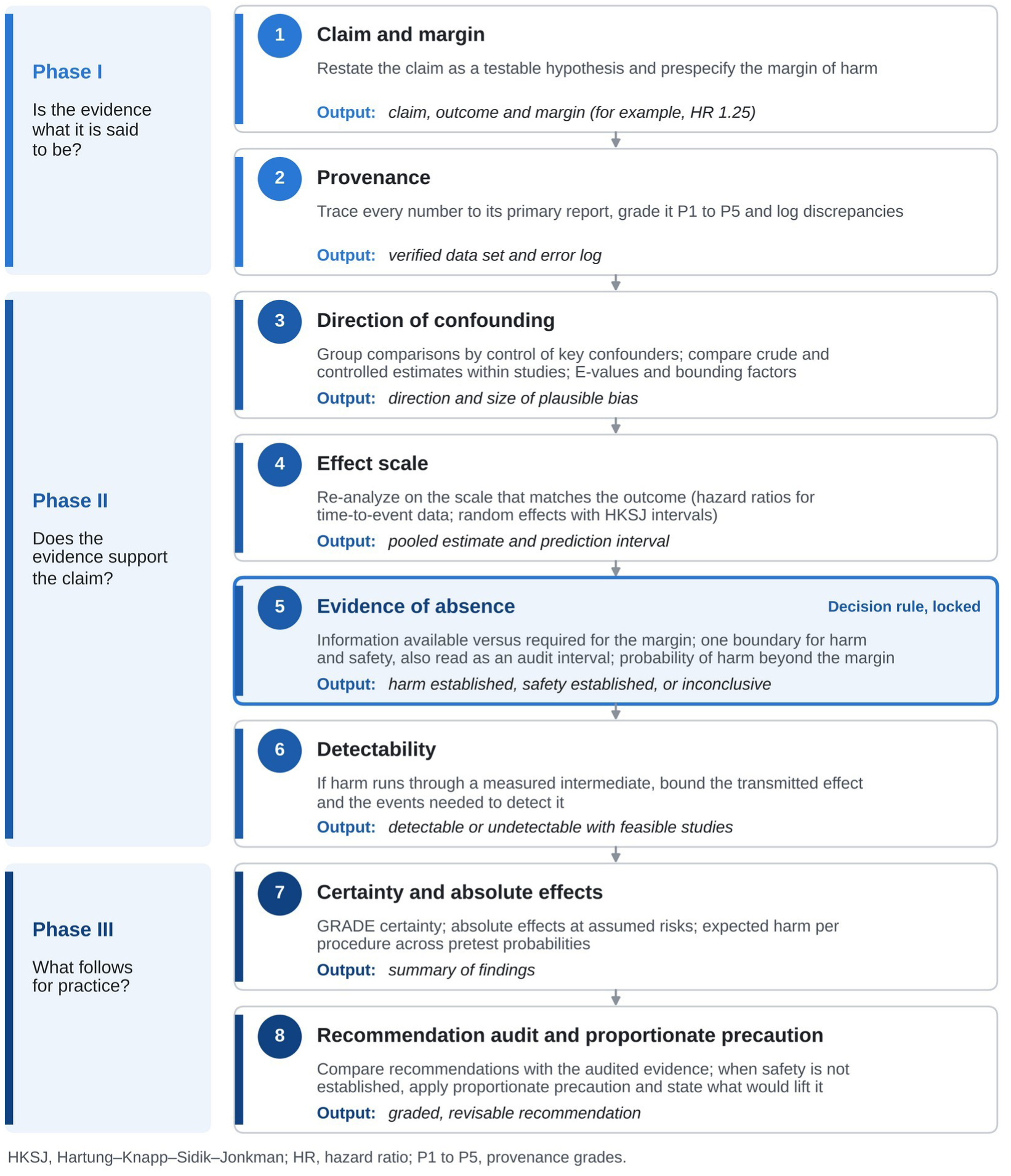
The eight-step framework for auditing claims of safety. Steps are grouped in three phases. Step 5 applies the same information-adjusted boundary to claims of harm and claims of safety. Formulas are given in S1 Appendix and a blank worksheet in S1 File. HKSJ, Hartung– Knapp–Sidik–Jonkman; HR, hazard ratio; P1 to P5, provenance grades.

**Table 1.** The eight-step framework for auditing a claim of safety.

| Step | Question for the reader | Methods | Output and decision rule |
|---|---|---|---|
| 1. Claim and margin | What harm, and of what size, is said not to occur? | Restate population, intervention, comparator and outcome; set the margin M before examining the evidence (here 1.25) | Testable claim; conclusions reported for alternative margins |
| 2. Provenance | Do the numbers match the original studies? | Trace every number to its primary report; grade P1 to P5; log discrepancies | P1 to P4 in primary analyses, P5 in sensitivity analyses only; discrepancies resolved in favor of the primary report |
| 3. Direction of confounding | Would the way patients were selected make the intervention look safer than it is? | Predict from the allocation mechanism; compare crude and controlled estimates within studies; group studies by a design feature; E-values and bounding factors | If controlled estimates move toward harm, crude comparisons are not accepted as evidence of safety |
| 4. Effect scale | Were recurrence and survival analyzed over time, or as simple proportions? | Hazard ratios for time-dependent outcomes, reported or reconstructed; random effects with HKSJ intervals; prediction intervals | Pooled cumulative proportions are not accepted as evidence of absence |
| 5. Evidence of absence | Was there enough information to detect the harm that matters, and was it excluded? | z<sub>harm</sub> and z<sub>safe</sub>; information fraction t = D / D<sub>req</sub>; boundary b(t) = 1.96 / √min(t, 1); audit interval HR × exp(±b(t)·SE); P(HR > 1) and P(HR > M) | Harm established if z<sub>harm</sub> ≥ b(t) (audit interval above 1); otherwise safety established if z<sub>safe</sub> ≥ b(t) (audit interval below M); otherwise inconclusive; can also be read from the two limits of a published 95% confidence interval, without software |
| 6. Detectability | Could the studies have detected the harm at all? | Bound the effect transmitted through an intermediate for k from 0 to 1; events needed to detect it | If the events needed far exceed those available, absence of an outcome signal is uninformative |
| 7. Certainty and absolute effects | How certain is the evidence, and what does it mean per 1,000 patients? | GRADE; absolute effects at assumed baseline risks; expected harm per procedure across pretest probabilities | Summary of findings with standardized statements; harm per 1,000 patients and per 1,000 procedures |
| 8. Recommendation audit and proportionate precaution | Do recommendations say what was excluded, and is any precaution proportionate and revisable? | Audit of role, grading and reliance on nonsignificance; knowledge and harm conditions; consistency and efficiency of candidate measures | Stratified, revisable recommendation, proportionate to the harm per 1,000 at stake, stating the evidence that would lift or strengthen it |

D = information in event-equivalents (4/SE<sup>2</sup> of the log hazard ratio); D<sub>req</sub> = information required to detect the margin with 80% power at a two-sided α of 0.05; GRADE = Grading of Recommendations Assessment, Development and Evaluation; HKSJ = Hartung–Knapp–Sidik–Jonkman; HR = hazard ratio; k = fraction of intervention-induced intermediate events carrying the prognostic weight of spontaneous ones; P1 to P5 = provenance grades (copied from full text, copied from abstract, derived arithmetically, reconstructed statistically, approximated); SE = standard error of the log hazard ratio.

#### Step 1. Claim and margin

The claim is restated as a testable proposition that specifies population, intervention, comparator and outcome, together with a margin M of clinically important harm on the relative scale. We used M = 1.25 for risk and hazard ratios, the GRADE threshold for appreciable harm [9]. The margin should be set by clinical judgment before the evidence is examined, and conclusions should be reported for alternative margins; here they are reported from 1.10 to 1.50.

#### Step 2. Provenance

Every number is traced to its primary report and graded: P1, copied from the full text; P2, copied from the abstract; P3, derived arithmetically from reported percentages and denominators; P4, reconstructed statistically, for example a hazard ratio from the log-rank P value and the number of events [34,35]; and P5, approximated, for example with events estimated from Kaplan–Meier proportions. Estimates graded P1 to P4 enter primary analyses and estimates graded P5 enter only sensitivity analyses. Discrepancies between secondary sources and primary reports are logged and resolved in favor of the primary report.

#### Step 3. Direction of confounding

For observational comparisons, the mechanism by which patients come to receive the intervention is used to predict the direction of confounding. The prediction is tested within studies, by comparing crude and controlled estimates of the same comparison, and between studies, by grouping comparisons according to whether the key prognostic factor was controlled by randomization, adjustment, stratification or restriction. The grouping variable must be a feature of study design, not of results. The strength of unmeasured confounding needed to mask harm at the margin is expressed as an E-value, and documented imbalances are translated into the Ding–VanderWeele bounding factor [15,36]. If controlled estimates consistently move toward harm, crude comparisons are not accepted as evidence of safety.

#### Step 4. Effect scale

Outcomes that depend on time, such as recurrence and survival, are analyzed as time-to-event data. Pooled cumulative proportions, which ignore censoring and unequal follow-up [34], are not accepted as evidence of absence; hazard ratios are taken as reported or reconstructed (Step 2). Estimates are pooled with random-effects models and Hartung–Knapp–Sidik–Jonkman (HKSJ) confidence intervals [37,38], and prediction intervals are reported [39].

#### Step 5. Evidence of absence

For an estimated hazard ratio HR, with standard error SE of its logarithm, two statistics are computed: z_harm_ = ln HR / SE, against the null hypothesis of no effect, and z_safe_ = (ln M – ln HR) / SE, against the null hypothesis that the effect equals the margin. Available information is expressed as event-equivalents, D = 4 / SE^2^, and compared with the information required to detect the margin with 80% power at a two-sided α of 0.05, D_req_ = 4(z_0.975_ + z_0.80_)^2^ / (ln M)^2^ [40], which is 631 event-equivalents for M = 1.25. When the standard error is recovered from a reported interval, the interval’s own critical value is used: 1.96 for a normal-based 95% confidence interval and the t quantile for an interval based on the t distribution, such as an HKSJ interval. With information fraction t = D / D_req_, both statistics are compared with the same O’Brien–Fleming-type boundary, b(t) = 1.96 / √min(t, 1), which for a single analysis is the Lan– DeMets boundary used in trial sequential analysis [12,41]. Harm is established if z_harm_ ≥ b(t); otherwise, safety (absence of harm beyond the margin) is established if z_safe_ ≥ b(t); otherwise the evidence is inconclusive. The rule is symmetric: claims of harm and claims of safety must meet the same standard. At full information it reduces to a conventional test of no effect and to a one-sided equivalence test at the margin [14]; below full information it guards against declaring either conclusion from small or early bodies of evidence.

The verdict can also be read from a single interval. The audit interval, HR × exp(±b(t)·SE), is the conventional 95% confidence interval widened on the logarithmic scale by the factor 1/√min(t, 1); it coincides with the confidence interval once information is complete and corresponds to the adjusted intervals of trial sequential analysis and to repeated confidence intervals in group sequential trials [12,42]. Harm is established when the lower limit of the audit interval is at least 1, safety when its upper limit is at most the margin, and otherwise the evidence is inconclusive. In plain terms, the audit interval is the range of effects that the evidence cannot yet rule out once its limited size is taken into account: the less information, the wider the range, so that with a quarter of the required information it is twice as wide, on the logarithmic scale, as the confidence interval. A claim of safety is accepted only if even the upper end of this range lies below the smallest harm that matters, and a claim of harm only if even its lower end lies above no effect. Probabilities that the hazard ratio exceeds 1 and M are reported alongside the verdict.

Because the verdict depends only on the two limits of a normal-based 95% confidence interval and on the margin, it can also be read from a published interval without software. Table 2 gives, for a margin of 1.25, the limits that establish safety or harm for each ratio of the upper to the lower limit, and states in plain terms what each verdict means and what follows from it.

**Table 2.**
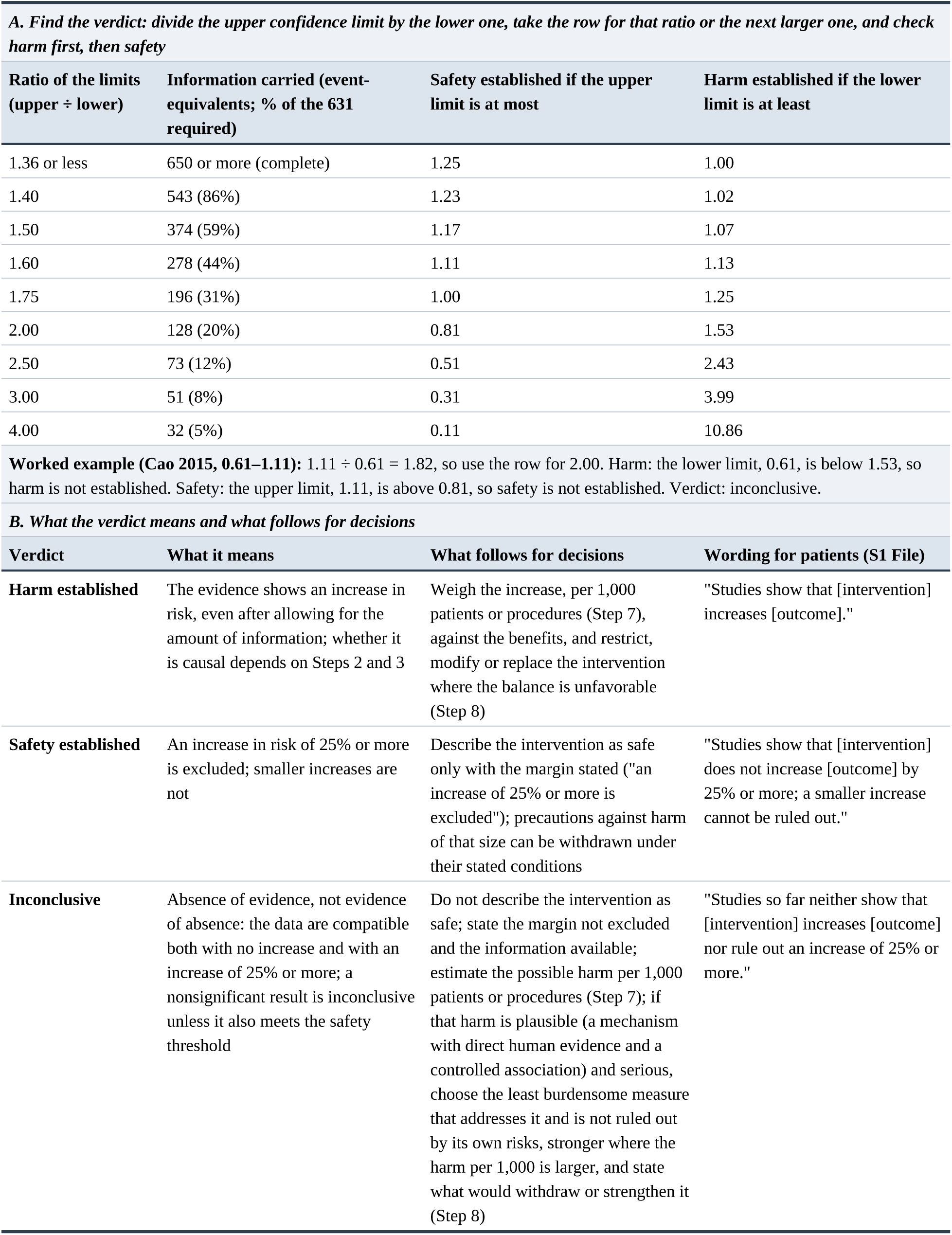
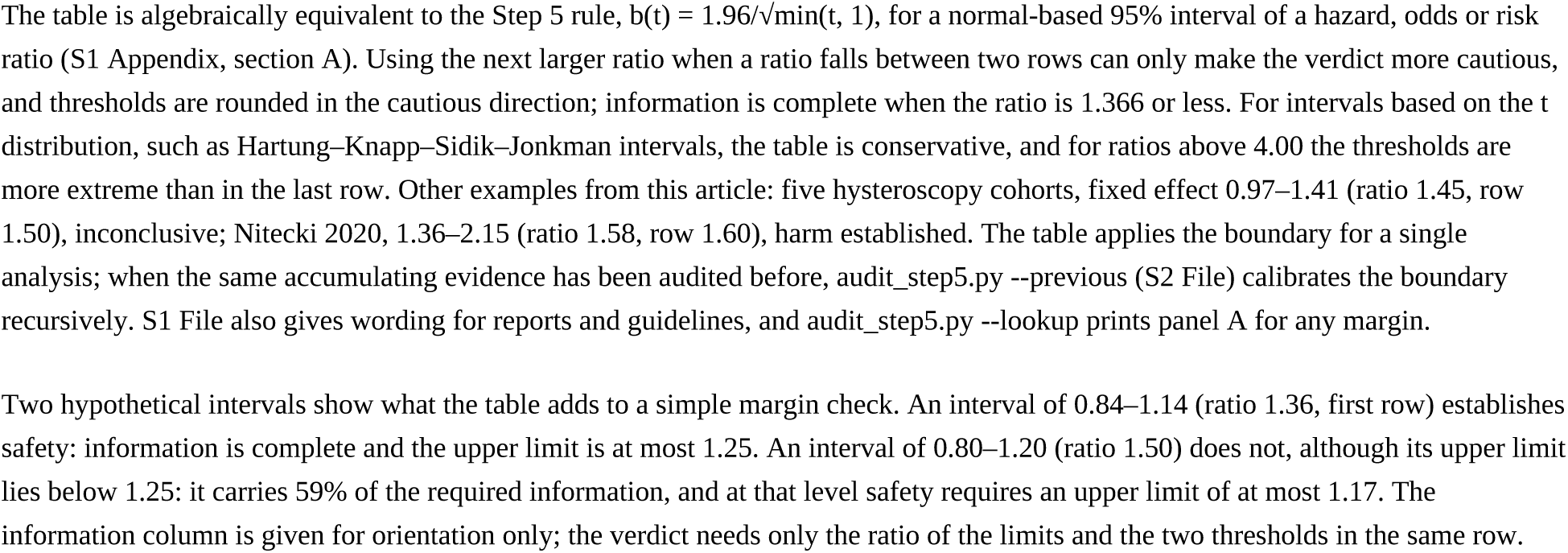
Reading the Step 5 verdict from a reported 95% confidence interval at a margin of 1.25, and what each verdict means for decisions.

#### Step 6. Detectability

When harm is hypothesized to act through a measured intermediate, the effect that the intermediate could transmit to the final outcome is bounded and compared with the available information [43]. The transmitted hazard ratio depends on the baseline risk of the intermediate, the effect of the intervention on it, the prognostic weight of the intermediate, and the fraction k of intervention-induced intermediate events that carry the same prognostic weight as spontaneous ones. Because k is rarely known, it is varied over its full range. If detecting the transmitted effect would require far more events than exist, the absence of a signal in the final outcome says little about the mechanism.

#### Step 7. Certainty and absolute effects

Certainty is rated with GRADE, in which observational evidence starts at low certainty [10], and findings are stated in standardized wording [44]. Relative effects are converted to absolute effects at assumed baseline risks, using p_1_ = OR·p_0_ / (1 – p_0_ + OR·p_0_) for odds ratios (ORs) and r_1_ = 1 – (1 – r_0_)^HR^ for hazard ratios [45]. When the intervention is used before the diagnosis is known, the expected harm per procedure is the absolute effect multiplied by the pretest probability of the disease. The two denominators answer different questions and are labeled throughout: per 1,000 patients who have the disease describes the harm to those affected, whereas per 1,000 procedures describes the harm across everyone examined, including the majority without the disease.

#### Step 8. Recommendation audit and proportionate precaution

Each relevant recommendation is examined for the role it assigns to the intervention, whether it addresses the harm in question, whether it grades that evidence, and whether it infers safety from nonsignificance. When Step 5 has not established safety, precaution is assessed with Steel’s conditions [19]. The knowledge condition is met when there is a plausible mechanism with direct human evidence and at least one controlled association with the outcome or with an intermediate of known prognostic weight; the harm condition is met when the potential harm is serious. Candidate measures are compared for consistency, rejecting any measure whose own risks would meet the same conditions [18], and for efficiency, preferring the least burdensome measure that addresses the risk. Recommendations are stratified by pretest probability and completeness of information, so that each measure is proportionate to the absolute harm at stake estimated in Step 7, and each states the evidence that would lift or strengthen it.

### Evaluation of the Step 5 rule

We compared three rules for declaring an intervention safe: rule 1, no statistically significant difference (two-sided P ≥ 0.05); rule 2, an upper 95% confidence limit below the margin, whatever the information; and rule 3, the Step 5 rule. For a single analysis, the probability of each verdict was computed exactly under normal theory for true hazard ratios of 1.0 and 1.25 and information from 15 to 4,000 event-equivalents. To represent a synthesis that is updated as studies accumulate, we simulated 200,000 bodies of evidence analyzed 10 times at equal increments of information up to the required information, using independent increments of the score statistic [41], and recorded the probability of declaring safety or harm at any update (S1 Appendix). Because the locked boundary does not adjust later analyses for earlier ones, we also calibrated boundaries recursively, by Monte Carlo simulation of 1,000,000 paths, so that the probability of first crossing at each update equals the α spent since the previous update under Lan–DeMets alpha spending [41].

### Retrospective test

To examine whether the framework would have distinguished absence of evidence from evidence of absence in a case whose answer is now known, we applied Steps 2 to 5 to minimally invasive radical hysterectomy for early-stage cervical cancer. Before 2018, observational studies and meta-analyses had concluded that the minimally invasive approach was oncologically safe [46,47]; a randomized trial then showed worse disease-free and overall survival [48,49]. The concern, exposure of tumor during manipulation, is related to that raised by hysteroscopy. We audited a large matched cohort whose authors concluded that laparoscopy was a safe alternative [46] and a pre-trial meta-analysis that reached the same conclusion [47] and, for comparison, the trial and two later meta-analyses [50,51]. The Step 5 rule had been locked before these data were analyzed, so the test could have failed: had the pre-trial evidence crossed the safety boundary, the framework would have endorsed a claim that the trial later contradicted. The selection of evidence is ours; the test demonstrates the behavior of the framework but cannot estimate its accuracy.

### Application to diagnostic hysteroscopy in endometrial cancer

#### Sources and eligibility

Primary comparative studies were identified from published meta-analyses [26,27,30,32,52,53], from tabulations in narrative reviews [54,55], and from targeted searches of web-indexed MEDLINE records and publisher and repository pages (search terms in S1 Appendix); the search was last updated on 7 November 2026. Studies were eligible if they compared diagnostic hysteroscopy with nonhysteroscopic sampling or with no hysteroscopy in women with endometrial cancer and reported positive peritoneal cytology or recurrence. Guidelines were the current versions issued by major oncology and gynecologic societies that address the diagnosis of endometrial cancer or hysteroscopic practice [56–59], together with the published National Comprehensive Cancer Network statement on cytology [60].

#### Statistical analysis

For cytology, each study contributed its best-controlled estimate, in the order randomization, adjustment or stratification for prognostic factors, restriction to early stage, and crude counts. Following Step 3, studies were grouped by whether disease extent had been controlled, because confounding by indication was the bias of principal concern [5]: hysteroscopy is used more when the diagnosis is uncertain and the tumor small. This grouping was defined by a design feature of each comparison, but during the analysis rather than in a protocol; the change from crude to controlled estimates within the same studies was therefore taken as the more direct test. ORs were pooled with random-effects models (restricted maximum likelihood) and HKSJ intervals, with the variance factor not allowed below 1 [37,38]; heterogeneity was summarized with I^2^ [61], and small-study effects were assessed with Egger’s test [62]. For recurrence, log hazard ratios for recurrence, progression or disease-free survival were pooled in the same way; the primary set contained estimates graded P1 to P4, and sensitivity sets added approximated estimates and crude recurrence ratios.

Probabilities that the hazard ratio exceeded 1 and 1.25 were obtained from the HKSJ t distribution and from Bayesian random-effects models with a reference prior (normal, mean 0, standard deviation 1 on the log scale) or a skeptical prior (5% probability that the hazard ratio exceeds 1.5), each with a half-normal prior (scale 0.5) on heterogeneity [63,64]. Cumulative fixed-effect z statistics were computed in chronological order for Step 5. For Step 6, the baseline risk of positive cytology was 4.4% for low-risk early-stage disease [28] and 10% for high-risk histology [65]; the effect of hysteroscopy was the extent-controlled pooled OR or, for high-risk histology, the high-risk stratum of Zerbe and colleagues [66]; the hazard ratio for positive cytology was 1.98 (95% CI 1.56–2.52) [28]; k was uniformly distributed between 0 and 1; and uncertainty was propagated with 200,000 Monte Carlo draws. For Step 7, the assumed 5-year recurrence risks without hysteroscopy were 5% and 30%, close to the control-group risks in an apparent early-stage cohort (4.7%) [67] and a type II cohort (32.6%) [65], and pretest probabilities were taken from common referral scenarios [22,23,68,69]. The sensitivity of the conclusions was examined for the margin (1.10 to 1.50), for the boundary (recursive calibration for the chronological updates), for k (0, 0.5 and 1) and for the pretest probability of cancer (1.9% to 42.6%).

### Data, code and software

Every result in this article can be regenerated with one command from the files in S2 File: the data set, with the provenance grade of every number, the code that reproduces every number, table and figure, and a script that compares a fresh run with the 381 reported numbers that the analysis produces, each listed with its location in the article. Analyses were run in Python 3.13.18 with NumPy 2.5.3, SciPy 1.18.1, Matplotlib 3.11.2 and Pillow 12.3.0; Monte Carlo analyses use fixed random seeds, so repeated runs give identical results. The same files allow the audit to be extended: a new study is added as one line of data and the whole analysis rerun. S2 File also contains a stand-alone script that applies the Step 5 rule to any ratio estimate and its confidence interval, returning the verdict and the audit interval, with recursively calibrated boundaries when the same evidence has been audited before, and that prints, for any margin, the confidence limits that establish safety or harm (Table 2 for a margin of 1.25). The same rule is implemented in a single-file calculator that runs in any web browser, offline and in English or Spanish: from an estimate and its confidence interval it gives the verdict with its audit interval, what the verdict means and what follows for decisions, how two common rules would read the same result, the absolute effects of Step 7, the conditions for precaution of Step 8 and the wording of S1 File, and a built-in self-check recomputes the examples of this article and compares them with the values reported here. S1 File is a worksheet for applying all eight steps without software, with a protocol for independent audits, a one-page committee summary, blank and completed for the hysteroscopy audit, wording for reports, guidelines and patients for each verdict, and a minimal check for journals, reviewers and guideline panels. None of these materials verifies the provenance of the numbers entered, which remains the user’s responsibility, and their output is not a clinical recommendation. The calculator is available at https://evidenceofharm.netlify.app/, and the data, code and calculator are archived at Zenodo (https://doi.org/10.5281/zenodo.24059401).

### Use of large language models

A large language model (Claude Opus 5 and Claude Opus 5.5; Anthropic) was used solely to assist with language editing, sentence-level polishing, and the organisation and formatting of the manuscript text. It was also used, in a final checking phase only, to help verify that the analytical code reproduced the quantities already obtained by the authors and to make the code and the calculator more robust (input checks, safeguards for extreme inputs and automated checks that the different implementations agree), without changing any reported result. Every numerical result reported in this paper was regenerated independently by the authors’ canonical analysis script and confirmed against the deposited reproducibility ledger. The model was not used to design the study, to analyse or interpret the data, to generate scientific content, or to draw conclusions. It does not meet authorship criteria and is not an author. All scientific content, analyses, interpretations and conclusions are the sole responsibility of the authors, who reviewed and take full responsibility for the final manuscript.

## Results

### Operating characteristics of the Step 5 rule

At the information available for hysteroscopy (449 event-equivalents), inferring safety from nonsignificance (rule 1) would have given false reassurance in 34% of analyses if the true hazard ratio were 1.25; the Step 5 rule did so in 1%. With the information of a small cohort (29 event-equivalents), the corresponding probabilities were 91% and less than 0.1%, and with 171 event-equivalents they were 69% and less than 0.1% (Fig 2A). Rule 2 held false reassurance at 2.5% for a single analysis, and the Step 5 rule never exceeded 2.5% at any level of information. When the true hazard ratio was 1.0, rule 1 declared safety in 95% of analyses whatever the information, whereas the Step 5 rule did so in 52% at 449 event-equivalents, 80% at 631 and 94% at 1,000 (Fig 2B): safety could be established, but only with adequate information. The probability of establishing harm when the true hazard ratio was 1.25 mirrored that of establishing safety when it was 1.0 (Fig 2C). All single-analysis probabilities are exact.

**Fig 2.**
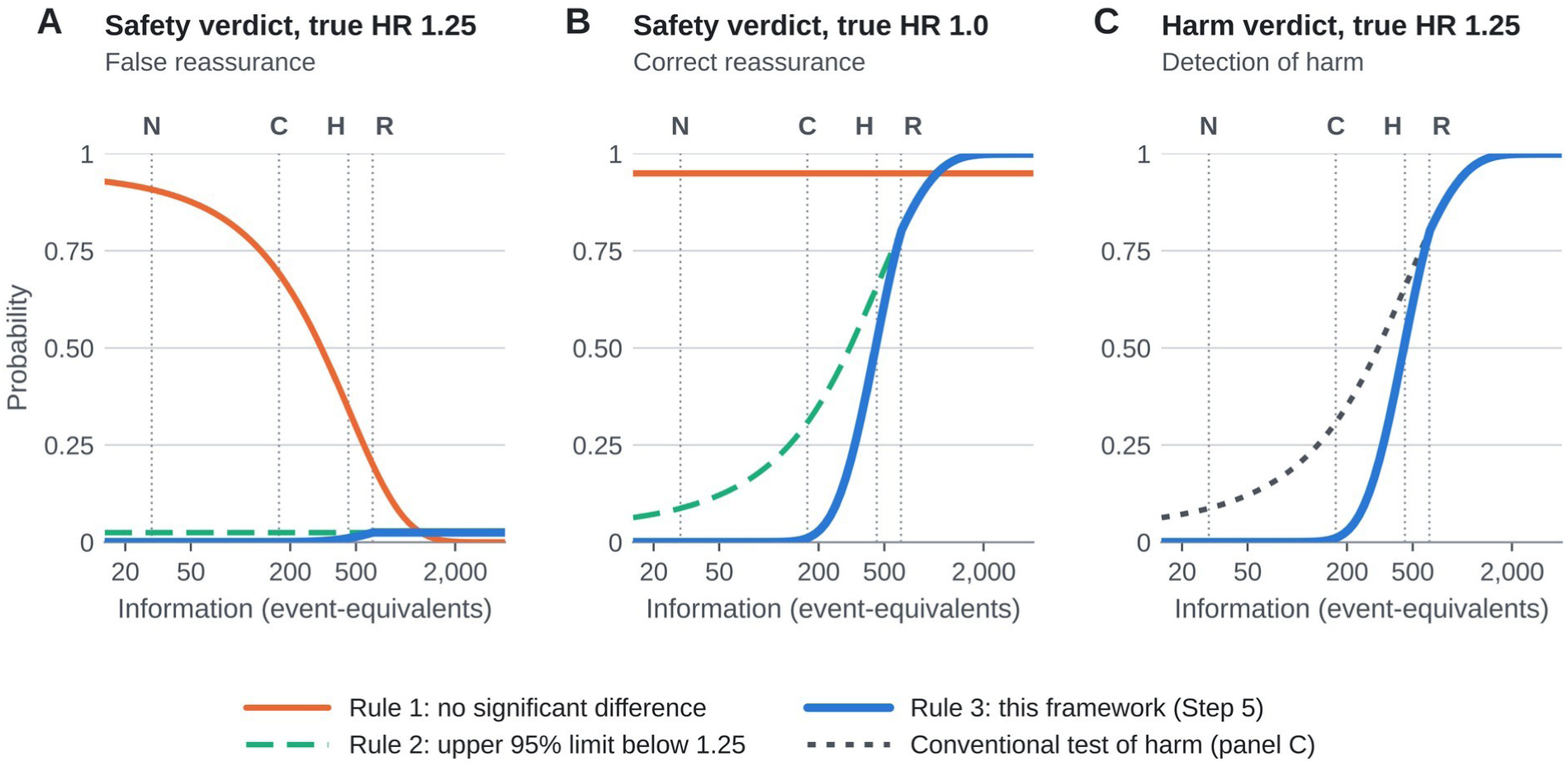
Operating characteristics of three rules for declaring an intervention safe. (A) Probability of a safety verdict when the true hazard ratio is 1.25, the margin of clinically important harm (false reassurance). (B) Probability of a safety verdict when the true hazard ratio is 1.0 (correct reassurance). (C) Probability of a harm verdict when the true hazard ratio is 1.25. Rule 1 infers safety from a nonsignificant test of no difference; rule 2 from an upper 95% confidence limit below 1.25; rule 3 is Step 5 of the framework. Probabilities are exact under normal theory for a single analysis. Vertical lines mark the information in the matched cohort of Nam and colleagues (N, 29 event-equivalents), in the pre-trial meta-analysis of Cao and colleagues taken at face value (C, 171), in the hysteroscopy cohorts combined (H, 449) and required for a margin of 1.25 (R, 631). HR, hazard ratio.

With 10 cumulative updates of the same evidence, repeated application of rule 2 declared safety at some update in 9.7% of simulated bodies of evidence (19,415 of 200,000) when the true hazard ratio was 1.25, and a conventional test declared harm at some update in 9.6% (19,233 of 200,000) when it was 1.0. With the locked Step 5 boundary these probabilities were 3.5% (6,916) and 3.4% (6,772), and the probability of eventually reaching the correct conclusion was 82% in both scenarios (164,102 and 163,979 of 200,000). Recursively calibrated boundaries, which fell from 6.20 at the first update to 2.12 at the last, reduced the two error probabilities to 2.5% (5,068) and 2.4% (4,895), at the cost of a lower probability of reaching the correct conclusion, 78% (155,677 and 155,479) (S1 Table).

### Retrospective test: minimally invasive radical hysterectomy

The matched cohort reported a hazard ratio for recurrence of 1.28 (0.62–2.64) [46]. It contained about 29 event-equivalents, 5% of the information required for a margin of 1.25; the probability of a hazard ratio above 1 was 75% and above 1.25 was 53%, and Step 5 classified the evidence as inconclusive (Table 3), with an audit interval of 0.04–37 that spans almost every plausible effect (Fig 3). The pre-trial meta-analysis pooled cumulative recurrence proportions from 13 studies (2,274 patients) with a fixed-effect model and reported an OR of 0.82 (0.61– 1.11) [47]. Its upper confidence limit lay below 1.25, so a margin check alone (rule 2) would have accepted the claim of safety. The framework would not have. The article labeled the same survival effects as hazard ratios in its text and as weighted mean differences in a table (Step 2).

**Fig 3.**
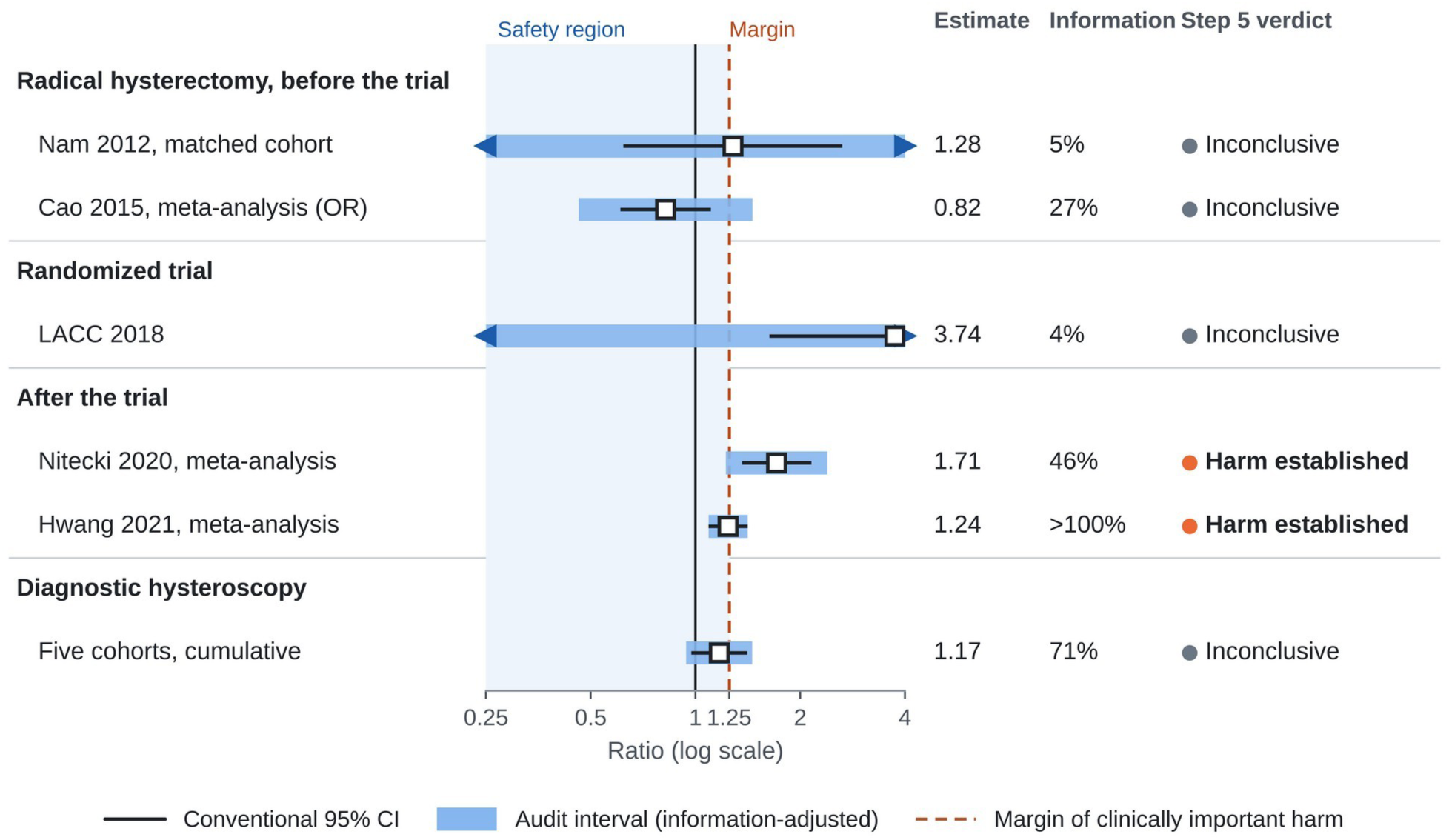
Step 5 read as an audit interval: the retrospective test and diagnostic hysteroscopy. For each body of evidence, the thin line is the conventional 95% confidence interval and the thick blue bar is the audit interval, the estimate multiplied by exp(±b[t]·SE), which widens the confidence interval on the logarithmic scale by the factor 1/√t when information is incomplete. Harm is established when the audit interval lies entirely above 1 (solid line), and safety when it lies entirely below the margin of 1.25 (dashed line; shaded region); otherwise the evidence is inconclusive; the dot beside each verdict repeats it (orange, harm established; gray, inconclusive). Arrows mark audit intervals that extend beyond the axis. Information is the percentage of the 631 event-equivalents required for a margin of 1.25. For Cao 2015, Step 5 was applied at face value to an odds ratio for cumulative recurrence proportions; the hysteroscopy row shows the cumulative fixed-effect estimate of the five cohorts with time-to-event data. CI, confidence interval; LACC, Laparoscopic Approach to Cervical Cancer trial; OR, odds ratio; SE, standard error of the log ratio.

**Table 3.** Retrospective test: evidence on minimally invasive radical hysterectomy for early-stage cervical cancer before and after the randomized trial.

| **Evidence** | **Design; patients** | **Estimate (95% CI)** | **Steps 2 to 4** | **Step 5: information; z vs boundary; audit interval** | **Verdict** |
|---|---|---|---|---|---|
| ***Before the trial (authors concluded that the minimally invasive approach was safe)*** |  |  |  |  |  |
| Nam 2012 [46] | Matched cohort; 526 | Recurrence HR 1.28 (0.62–2.64) | Time-to-event; matched design | 29 (5%); z<sub>harm</sub> 0.67, z<sub>safe</sub> −0.06 vs 9.1; 0.04–37 | Inconclusive |
| Cao 2015 [47] | Meta-analysis of 13 observational studies; 2,274 | Recurrence OR 0.82 (0.61–1.11), fixed effect | Step 2: effects labeled inconsistently; Step 3: unadjusted observational comparisons; Step 4: cumulative proportions | 171 (27%) at face value; z<sub>safe</sub> 2.76 vs 3.76; 0.46–1.46 | Inconclusive; fails Steps 2 to 4 |
| ***Randomized trial*** |  |  |  |  |  |
| LACC 2018 [48] | Randomized trial; 631 | Recurrence or death HR 3.74 (1.63–8.58) | Randomized; time-to-event | 22 (4%); z<sub>harm</sub> 3.11 vs 10.4; 0.05–310; P(HR > 1.25) >99% | Inconclusive by Step 5; knowledge condition for precaution met |
| ***After the trial*** |  |  |  |  |  |
| Nitecki 2020 [50] | Meta-analysis of 15 observational studies controlling for tumor size or stage; 9,499 | Recurrence or death HR 1.71 (1.36–2.15) | Controlled; time-to-event | 293 (46%); z<sub>harm</sub> 4.59 vs 2.88; 1.22–2.39 | Harm established |
| Hwang 2021 [51] | Meta-analysis of 36 studies | DFS HR 1.24 (1.09–1.41) | Step 2: larger with reported HRs, 1.41 (1.21–1.64); Step 3: larger in matched designs, 1.49 (1.19–1.88) | 928 (>100%); z<sub>harm</sub> 3.28 vs 1.96; 1.09–1.41 | Harm established |

Information in event-equivalents (4/SE<sup>2</sup> of the log ratio) and, in parentheses, as a percentage of the 631 required for a margin of 1.25; boundary b(t) = 1.96/√min(t, 1); audit interval = estimate × exp(±b(t)·SE). For Cao 2015, Step 5 was applied at face value, treating the odds ratio for cumulative recurrence proportions as if it were a hazard ratio. CI = confidence interval; DFS = disease-free survival; HR = hazard ratio; LACC = Laparoscopic Approach to Cervical Cancer trial; OR = odds ratio; SE = standard error.

Step 3 predicts confounding toward safety if the minimally invasive approach was used preferentially for smaller tumors, a concern that later syntheses addressed by restricting to studies that controlled for tumor size or stage [50]. Cumulative proportions ignore censoring and unequal follow-up (Step 4). And even taken at face value, the estimate contained 171 event-equivalents, 27% of the information required, with z_safe_ of 2.76 against a boundary of 3.76; its audit interval, 0.46–1.46, extended beyond the margin (Step 5: inconclusive).

The randomized trial reported a hazard ratio for recurrence or death from cervical cancer of 3.74 (1.63–8.58) [48]. With about 22 event-equivalents (4% of the information required), this single trial did not cross the Step 5 boundary (z_harm_ 3.11 against 10.4; audit interval 0.05–310), although the probability of harm beyond 1.25 exceeded 99%; such evidence meets the knowledge condition for precaution in Step 8, and enrollment had been stopped before the planned sample size was reached, after a safety alert from the trial’s data and safety monitoring committee. Harm was established in the sense of Step 5 by later syntheses: 15 observational studies that controlled for tumor size or stage gave a hazard ratio of 1.71 (1.36–2.15), with z_harm_ 4.59 against a boundary of 2.88 and an audit interval of 1.22–2.39 [50], and a meta-analysis of 36 studies gave 1.24 (1.09–1.41), with z_harm_ 3.28 against 1.96 [51]. In the latter, estimates were larger when hazard ratios were taken as reported rather than reconstructed, 1.41 (1.21–1.64), and in matched designs, 1.49 (1.19–1.88), the pattern that Steps 2 and 3 anticipate. Because the rule had been locked beforehand, the classification of the pre-trial evidence as inconclusive was a genuine test result, not a property built into the analysis.

### Application to diagnostic hysteroscopy in endometrial cancer

#### Steps 1 and 2: claim, margin and provenance

The audited claim was that diagnostic hysteroscopy before surgery does not increase recurrence in women with endometrial cancer, with a margin of 1.25 and positive peritoneal cytology as the hypothesized intermediate. Of 29 comparative reports assessed, 18 contributed data and 11 could not be pooled (PRISMA flow diagram in S1 Fig; S2 Table; S3 Table).

Cytology data came from 12 studies with 4,078 patients, including one randomized trial [65,66,70–79], and recurrence data from 9 studies [31,65,67,76,77,80–83], 5 of them in the primary time-to-event set. The search update of 7 November 2026 added two reports: one contributed cytology data [79], and one could not be pooled because it compared operative hysteroscopy with a group that included diagnostic hysteroscopy and reported no hazard ratio [84]. Of the 17 study-level estimates in the primary analyses, 12 were copied from primary reports, 3 were derived arithmetically and 2 were reconstructed statistically. All analyses are summarized in S4 Table.

Several secondary tabulations differed from the primary reports in ways that can propagate into later syntheses (Table 4). In one review table, a randomized trial appeared as 8 of 38 patients with positive cytology, figures that match the number randomized to, and the number excluded from, its hysteroscopy arm, rather than the 3 of 30 reported; another review listed the same trial as 1 of 30 [54,55,71]. A flow rate of 150 mL/min was tabulated as an intrauterine pressure of 150 mmHg [85], and a cohort that included women without hysteroscopy was tabulated as 113 hysteroscopy patients [86]. Primary reports were not always internally consistent either: the abstract of the largest type II cohort assigns 269 patients to hysteroscopy and 160 to curettage, whereas its results and tables give 160 and 269, the figures that a 2024 meta-analysis used [53,82]. This is why provenance grading distinguishes numbers copied from the full text (P1) from those copied from the abstract (P2).

**Table 4.** Discrepancies found by tracing numbers to the primary reports (Step 2)

| Secondary source | Primary study | As tabulated | As reported in the primary study | Consequence |
|---|---|---|---|---|
| Review table, 2010 [54] | Sáinz de la Cuesta 2004, randomized [71] | 8/38 (21%) positive cytology | 3/30 (10%) vs 1/20 (5%); 38 randomized to and 8 excluded from the hysteroscopy arm | Effect in a randomized trial overstated |
| Review table, 2007 [55] | Same trial | 1/30 (10%) | 3/30 (10%) hysteroscopy; 1/20 control | Arms mixed; count and percentage inconsistent |
| Review table, 2010 [54] | Selvaggi 2003 [85] | 9/39 (23%); pressure 150 mmHg | 9 positive among all 147 women, not associated with procedure; pressure 25–50 mmHg, flow 150 mL/min | Unit error; low-pressure study shown as high pressure |
| Review table, 2010 [54] | Obermair 2000 [86] | 113 hysteroscopy patients, 10 positive | 113 women with or without hysteroscopy | Comparator counted as exposed |
| Review table, 2010 [54] | Kudela 2001 [87] | 4/134 (5.3%) | 4/75 with fluid obtained (5.3%), plus 8 suspicious | Denominator inflated; suspicious results omitted |
| Review table, 2010 [54] | Arikan 2001 [25] | 20/24 (83%) positive cytology | Transtubal fluid passage 20/24; tumor cells 17; viable cells 10 | Fluid passage recorded as cytology |
| Abstract, 2022 [82] | Zhou 2022, type II [82] | 269 hysteroscopy vs 160 curettage | 160 vs 269 in the results and tables, the figures used by a 2024 meta-analysis [53] | Arms reversed in the abstract; full text (P1) preferred over abstract (P2) |
| Meta-analysis, 2021 [30] | Pooled survival | Overall survival P = 0.02 and disease-specific survival P = 0.00 beside pooled effects | Heterogeneity tests (I<sup>2</sup> 80% and 93%); pooled effects not significant | Heterogeneity P values readable as effect P values |

Primary values as printed in the primary publications. The list illustrates types of discrepancy and is not exhaustive.

#### Step 3: direction of confounding

When disease extent was controlled, hysteroscopy was associated with positive cytology (6 studies; OR 2.86, 95% CI 1.24–6.62; I^2^ 32%) (Fig 4). In crude all-stage comparisons there was no association and heterogeneity was substantial (6 studies; OR 0.90, 95% CI 0.25–3.20; I^2^ 64%); the ratio of the two pooled ORs was 3.18 (95% CI 1.05–9.62; P = 0.04). The predicted direction was reproduced within studies: controlling for disease extent changed the OR from 2.11 to 3.88 [72], from 3.20 to 7.08 [65] and from 1.11 to 4.19 [75]. In cohorts without such control, hysteroscopy groups had less advanced disease (stage III–IV, 30.1% vs 43.8%) [77], or advanced stage and vascular invasion were more frequent after curettage [78], the pattern expected under confounding by indication. In the only cohort that reported both, adjustment also raised the hazard ratio for disease-free survival, from 1.25 to 1.38 [82].

**Fig 4.**
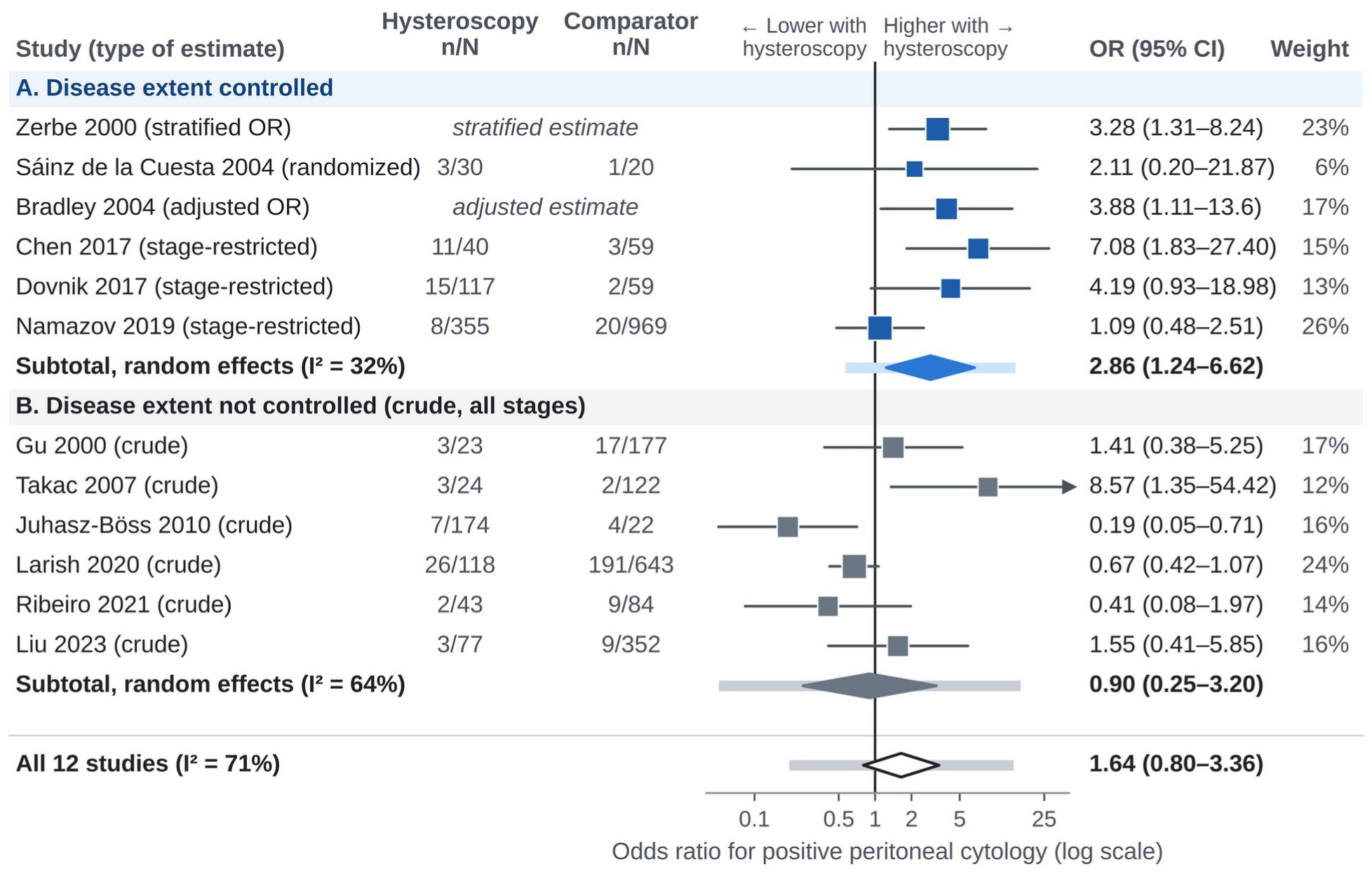
Positive peritoneal cytology after diagnostic hysteroscopy, by whether disease extent was controlled (Step 3). Each study contributes its best-controlled estimate (randomized comparison, adjusted or stratified OR, or restriction to early stage). Squares are proportional to random-effects weights within each subgroup; diamonds show pooled ORs with Hartung– Knapp–Sidik–Jonkman 95% CIs, and light bars show 95% prediction intervals; blue marks comparisons that controlled disease extent and gray those that did not. Ratio of subgroup Ors 3.18 (95% CI 1.05–9.62; P = 0.04). Zerbe 2000 combines two strata; counts for Dovnik 2017 (stage I) and Namazov 2019 were derived from reported percentages. CI, confidence interval; OR, odds ratio.

The extent-controlled estimate was robust to excluding studies with derived counts (OR 3.91, 95% CI 1.41–10.82) and to omitting any single study (2.41 to 3.95). The Bayesian posterior OR was 2.69 (95% credible interval 1.47–5.09), with a probability above 99% that the OR exceeds 1. Egger’s test did not suggest small-study effects (P = 0.12; S2 Fig), and published subgroups followed a gradient with pressure and medium: OR 1.18 (0.50–2.79) below 80 mmHg and 2.18 (1.28–3.73) above it [27], and 11.2 (1.3–94.5) for saline versus carbon dioxide [88] (S3 Fig). To mask a true hazard ratio of 1.25 for recurrence, unmeasured confounding would need an E-value of 1.81 if the observed estimate were 1.0, and of 2.19 for a pooled risk ratio of 0.88 [32]; the stage imbalance in one cohort [77], combined with a hazard ratio of 4.67 for advanced stage [78], yields a bounding factor of 1.33, enough by itself to make a true hazard ratio of 1.25 appear null.

#### Step 4: effect scale

Five cohorts provided time-to-event estimates (Fig 5). The pooled hazard ratio for recurrence or progression was 1.17 (95% CI 0.90–1.52), without detectable heterogeneity (I^2^ 0%; prediction interval 0.87–1.58). Adding approximated events gave 1.11 (0.91–1.35) and adding crude recurrence ratios gave 1.10 (0.92–1.32), moving the estimate toward 1 without excluding 1.25; omitting any cohort gave 1.12 to 1.24, and the hazard ratio for overall or disease-specific survival was 1.09 (0.67–1.78). Published syntheses that pooled cumulative recurrence proportions had reported risk ratios of 0.80 and 0.88, the latter with a fixed-effect model [30,32] (Fig 6); under Step 4, these estimates cannot serve as evidence of absence. The only cohort that reported a statistically significant adverse association, in advanced-stage disease, could not be pooled because its accessible report gave no confidence interval [89].

**Fig 5.**
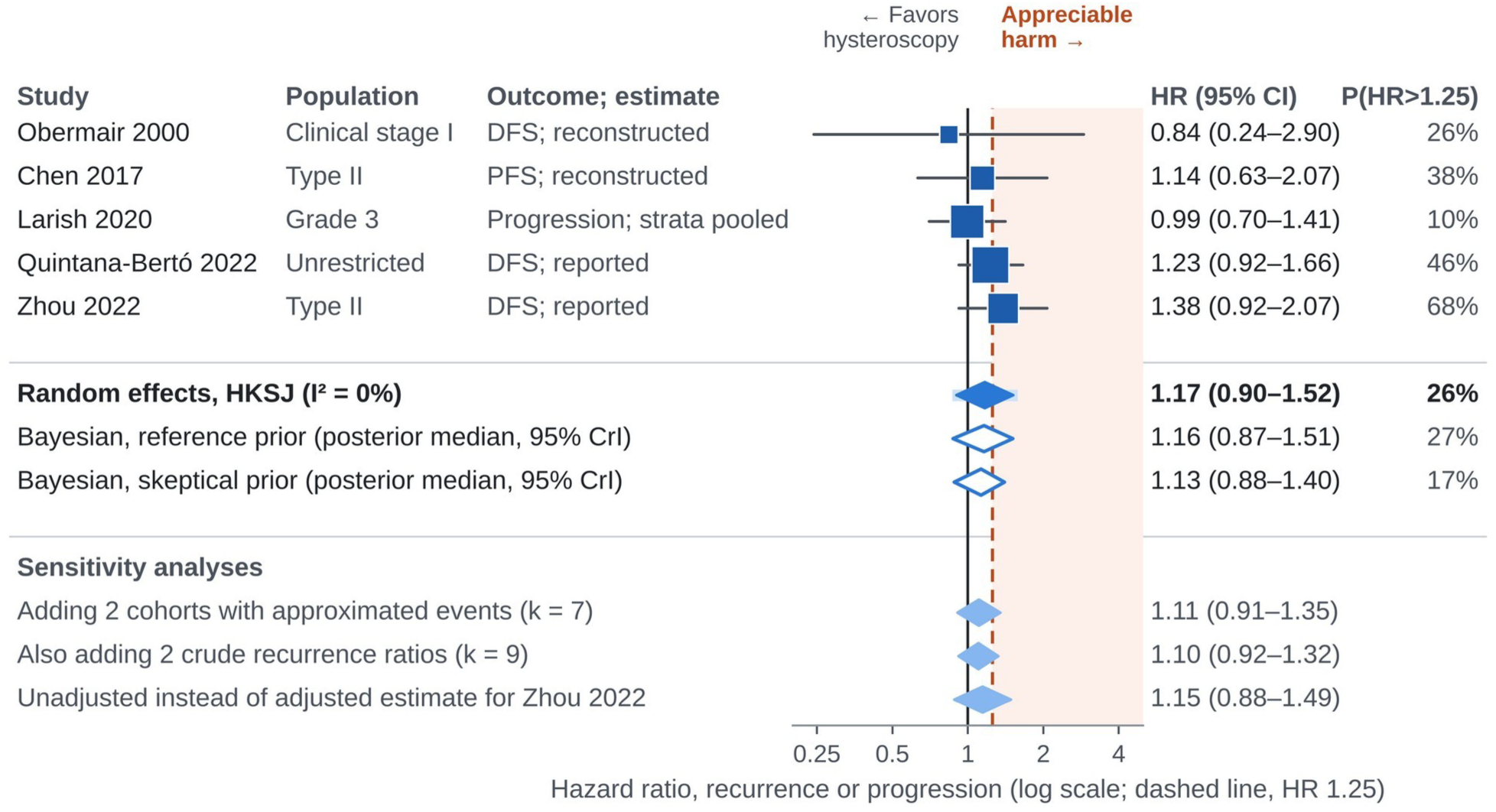
Recurrence or progression after diagnostic hysteroscopy: time-to-event estimates (Step 4). The shaded area marks hazard ratios above 1.25 (dashed line), the margin of clinically important harm. P(HR>1.25) is the probability that the hazard ratio exceeds 1.25 (flat prior for single studies, Hartung–Knapp t distribution for the pooled estimate, posterior probability for Bayesian rows). Reconstructed estimates were derived from the log-rank P value and number of events; Larish 2020 pools stage-specific estimates. Open diamonds show Bayesian estimates and light-blue diamonds sensitivity analyses. CrI, credible interval; DFS, disease-free survival; HKSJ, Hartung–Knapp–Sidik–Jonkman; HR, hazard ratio; PFS, progression-free survival.

**Fig 6.**
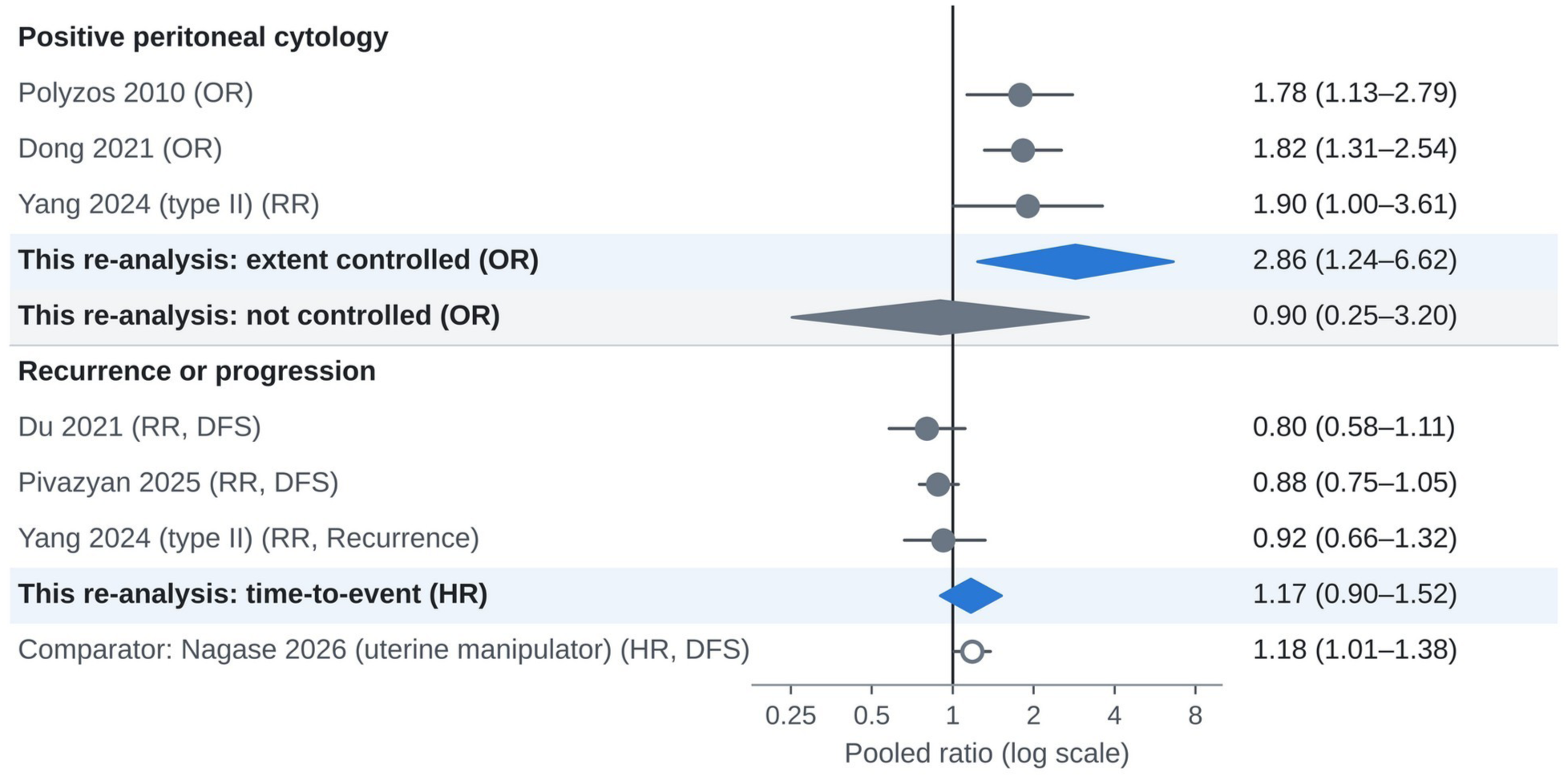
Published syntheses compared with this re-analysis (Step 4). Pooled estimates reported by published meta-analyses [26,27,30,32,53] (circles, with 95% CIs) for positive peritoneal cytology and for recurrence or progression, beside the estimates of this re-analysis (diamonds; blue, disease extent controlled or time-to-event analysis; gray, disease extent not controlled). The published estimates for recurrence are risk ratios of cumulative proportions, which under Step 4 cannot serve as evidence of absence; this re-analysis pooled hazard ratios. The open circle shows, for comparison, the hazard ratio for disease-free survival with uterine manipulators [4]. CI, confidence interval; DFS, disease-free survival; HR, hazard ratio; OR, odds ratio; RR, risk ratio.

#### Step 5: evidence of absence

The cohorts provided 449 event-equivalents, 71% of the information required for a margin of 1.25 and 48% of that for 1.20 (Fig 7). Information was computed from the standard error of the pooled log hazard ratio (S1 Appendix); recovering that standard error from the HKSJ interval with the normal quantile 1.96, rather than the t quantile for 4 degrees of freedom, would halve the apparent information. The cumulative z statistic was 1.65 for harm and 0.71 for safety, against a boundary of 2.32; neither crossed it, so the evidence was inconclusive, and the audit interval, 0.94–1.46, contained both 1 and the margin (Fig 3). The verdict was the same with boundaries calibrated recursively for the five chronological updates (final boundary 2.39). The probability that hysteroscopy increases the hazard was 91%, and that the increase exceeds 1.25 was 26% (Bayesian: 27% with the reference prior and 17% with the skeptical prior; S4 Fig; S5 Table). Had the true hazard ratio been 1.0, z_safe_ would have been expected to be about 2.36 with this information, and the Step 5 rule would have established safety in about half of analyses (Fig 2B); the observed value, 0.71, was well below that expectation. The verdict was the same at every margin from 1.10 to 1.40 (Table 5). Only for a margin of 1.50 did z_safe_ cross its boundary (2.64 against 1.96): a 50% or greater increase in the hazard of recurrence was excluded, although narrowly, because the random-effects interval reached 1.52. In clinical terms, the evidence makes a large increase in recurrence unlikely but cannot exclude the moderate increases that would matter to patients.

**Fig 7.**
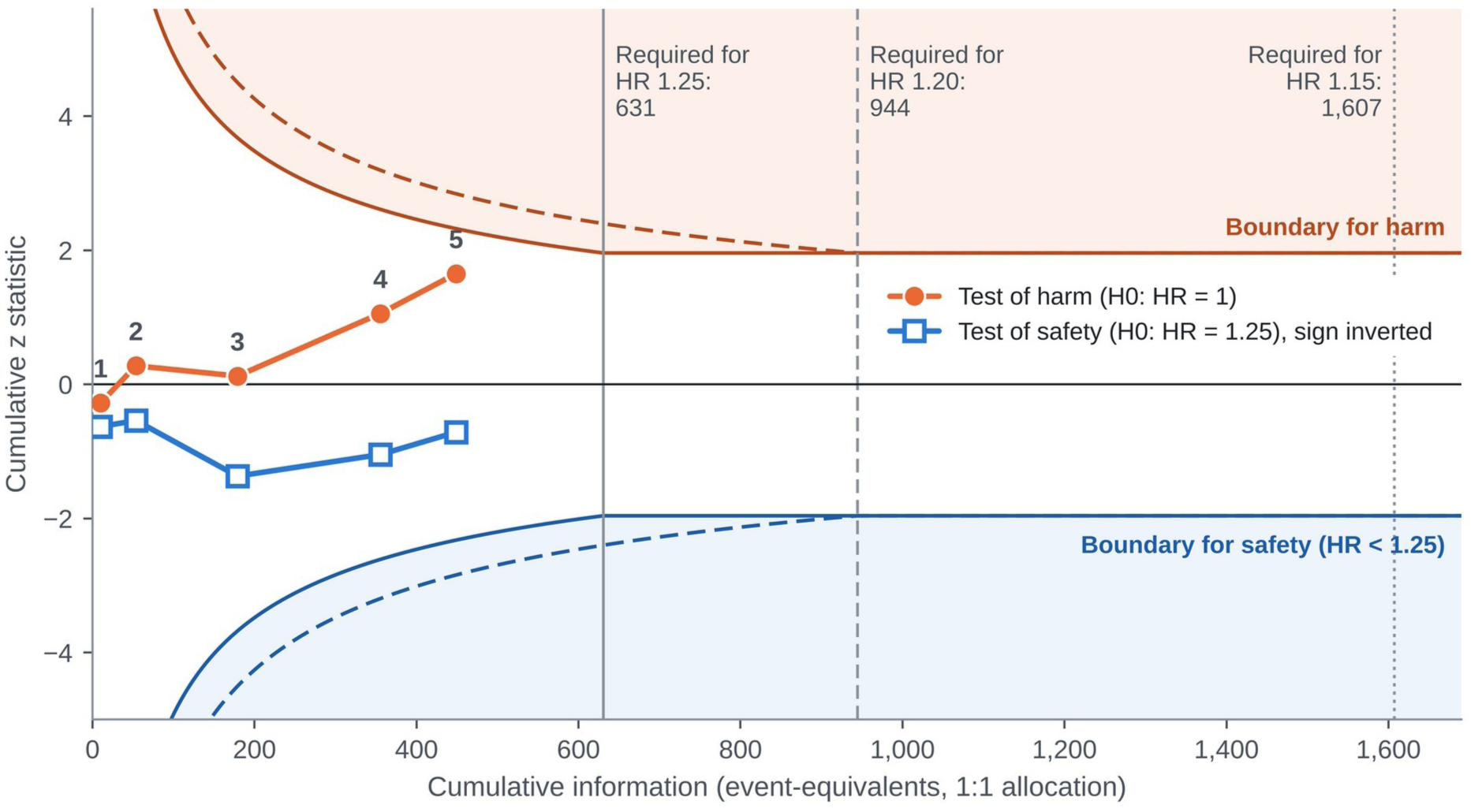
Cumulative information on recurrence (Step 5). Cumulative fixed-effect z statistics for the five cohorts with time-to-event data, added in chronological order (numbered points), for a test of harm (null hypothesis HR = 1; circles) and a test of safety (null hypothesis HR = 1.25; squares, plotted with inverted sign). Curves show the O’Brien–Fleming-type boundaries, b(t) = 1.96/√min(t, 1), for margins of 1.25 (solid) and 1.20 (dashed); harm or safety would be established only if the corresponding statistic crossed its boundary (shaded regions, margin 1.25). Vertical lines mark the information required for margins of 1.25 (631 event-equivalents), 1.20 (944) and 1.15 (1,607). With 449 event-equivalents, the cumulative z statistics were 1.65 for harm and 0.71 for safety, against a boundary of 2.32, so the evidence was inconclusive. H0, null hypothesis; HR, hazard ratio.

**Table 5.** Sensitivity of the conclusions for diagnostic hysteroscopy to the margin, the boundary, the prognostic weight of induced cytology and the pretest probability of cancer.

| Assumption varied | Value | Result | Implication |
|---|---|---|---|
| ***Margin of clinically important harm (Step 5; observed hazard ratio 1.17, 449 event-equivalents)*** |  |  |  |
| Margin | 1.10 | 13% of the 3,456 event-equivalents required; z<sub>safe</sub> −0.64 vs 5.44 | Inconclusive |
| Margin | 1.15 | 28% of 1,607; z<sub>safe</sub> −0.17 vs 3.71 | Inconclusive |
| Margin | 1.20 | 48% of 944; z<sub>safe</sub> 0.28 vs 2.84 | Inconclusive |
| Margin (main analysis) | 1.25 | 71% of 631; z<sub>safe</sub> 0.71 vs 2.32 | Inconclusive |
| Margin | 1.40 | Information complete; z<sub>safe</sub> 1.91 vs 1.96 | Inconclusive |
| Margin | 1.50 | Information complete; z<sub>safe</sub> 2.64 vs 1.96; random-effects upper limit 1.52 | Increase of 50% or more excluded, narrowly |
| ***Boundary (Step 5; margin 1.25)*** |  |  |  |
| Boundary | Recursive | Calibrated for the five chronological updates: final boundary 2.39 vs 2.32 locked; z<sub>harm</sub> 1.65, z<sub>safe</sub> 0.71 | Inconclusive; verdict unchanged |
| ***Prognostic weight of hysteroscopy-induced positive cytology, k (Step 6; excess recurrences per 1,000 women with cancer)*** |  |  |  |
| k | 0 | Transmitted hazard ratio 1.00 in low-risk and high-risk disease; no excess recurrence | Cytology pathway harmless; precaution would rest only on the direct time-to-event estimate |
| k | 0.5 | Low risk: HR 1.03, 1.7 more, about 28,100 events to detect. High risk: HR 1.08, 20 more, about 5,400 events | Small in low-risk disease; relevant in high-risk histology |
| k | 1 | Low risk: HR 1.07, 3.3 more, about 7,300 events. High risk: HR 1.16, 38 more, about 1,400 events | Clinically important in high-risk histology; undetectable by current cohorts |
| ***Pretest probability of cancer (Steps 7 and 8; per 1,000 procedures)*** |  |  |  |
| Asymptomatic endometrial polyp | 1.9% | 1.4 more positive cytology; 0.2 (up to 0.5) more recurrences if the pooled hazard ratio applied | Hysteroscopy as usual, at the lowest effective pressure, recorded |
| Symptomatic endometrial polyp | 5.1% | 3.7; 0.4 (up to 1.3) | As above |
| Postmenopausal bleeding | 9.0% | 6.5; 0.7 (up to 2.2) | As above, unless a focal lesion or other findings raise the probability |
| Bleeding and polyp, age 70 years or older | 13.1% | 9.5; 1.1 (up to 3.3) | Curettage first when preoperative information is incomplete |
| Atypical hyperplasia on endometrial sampling | 42.6% | 31; 3.5 (up to 11) | Curettage first when preoperative information is incomplete |

Margins: the information required is that needed to detect the margin with 80% power at a two-sided α of 0.05; boundary b(t) = 1.96/√min(t, 1). Recursive boundary: Lan–DeMets alpha spending calibrated by Monte Carlo simulation for the information at each chronological update. k: transmitted hazard ratios at the central inputs of Step 6 (low risk: positive cytology risk 4.4%, OR 2.86; high risk: 10%, OR 3.46), with 5-year recurrence risks of 5% and 30%. Pretest probabilities and sources as in S6 Table, where confidence intervals are given. HR = hazard ratio; OR = odds ratio.

**Table 6.** Summary of findings: diagnostic hysteroscopy compared with no hysteroscopy in women with endometrial cancer (Step 7)

| **Outcome and setting** | **Participants (studies)** | **Relative effect (95% CI)** | **Risk without HSC, per 1,000<sup>a</sup>** | **Difference with HSC, per 1,000 (95% CI)** | **Certainty (GRADE)<sup>b</sup>** | **What the evidence says<sup>c</sup>** |
|---|---|---|---|---|---|---|
| ***Positive peritoneal cytology, disease extent controlled*** |  |  |  |  |  |  |
| Low-risk early stage | 2,127 (6 studies, 1 randomized) | OR 2.86 (1.24–6.62) | 44 | 72 more (10 to 190 more) | Low | Hysteroscopy may increase positive cytology |
| High-risk histology |  |  | 100 | 141 more (21 to 324 more) |  |  |
| ***Recurrence or progression at 5 years*** |  |  |  |  |  |  |
| Low-risk early stage | 3,393 (5 cohorts) | HR 1.17 (0.90–1.52) | 50 | 8 more (5 fewer to 25 more) | Very low | The effect is very uncertain; harm beyond an HR of 1.25 is not excluded |
| High-risk histology |  |  | 300 | 41 more (26 fewer to 118 more) |  |  |
| ***Overall or disease-specific survival*** |  |  |  |  |  |  |
| All settings | 2,300 (3 cohorts) | HR 1.09 (0.67–1.78) | — | Not estimated | Very low | The effect is very uncertain |

<sup>a</sup>Assumed risks without hysteroscopy: positive cytology 4.4% in stage I endometrioid cancer [28] and 10% in high-risk histology [65]; 5-year recurrence 5% and 30%, close to the control-group risks in an apparent early-stage cohort (4.7%) [67] and a type II cohort (32.6%) [65]. Differences for recurrence were calculated from the hazard ratio [45]. All differences are per 1,000 women who have endometrial cancer; per 1,000 procedures they are obtained by multiplying by the pretest probability of cancer (Table 5; S6 Table). <sup>b</sup>Observational evidence starts at low certainty [10]. Positive cytology was not rated down for risk of bias, because residual confounding by indication would bias toward no effect, nor rated up, because the interval is wide and rests on few events. Recurrence and survival were rated down for imprecision: the intervals include no effect and appreciable harm, and the information available for recurrence is 48% of that required to detect an HR of 1.20. <sup>c</sup>Standardized GRADE wording [44]. CI = confidence interval; GRADE = Grading of Recommendations Assessment, Development and Evaluation; HR = hazard ratio; HSC = hysteroscopy; OR = odds ratio.

#### Step 6: detectability

In low-risk early-stage disease, the hazard ratio transmitted through cytology was small (median 1.03; 95% interval 1.00–1.12) and no higher than 1.07 even if every hysteroscopy-induced positive cytology carried full prognostic weight (Fig 8). Detecting a hazard ratio of that size would require about 7,300 to 50,000 events, 16 to 111 times the information now available. In high-risk histology, the transmitted hazard ratio reached 1.16 at full prognostic weight (median 1.06; 95% interval 1.00–1.27; probability above 1.10, 31%), detectable with about 1,400 events. The absence of a survival signal is therefore uninformative about the cytology pathway in low-risk disease and only weakly informative in high-risk disease. At the extremes of k the conclusions diverge (Table 5): if displaced cells were prognostically inert (k = 0), hysteroscopy would cause no survival harm through cytology and precaution would rest only on the direct time-to-event estimate; if they behaved like spontaneous peritoneal spread (k = 1), the excess would be 3.3 recurrences per 1,000 women with low-risk disease and 38 per 1,000 with high-risk histology, a clinically important harm that would still need about 1,400 events to detect.

**Fig 8.**
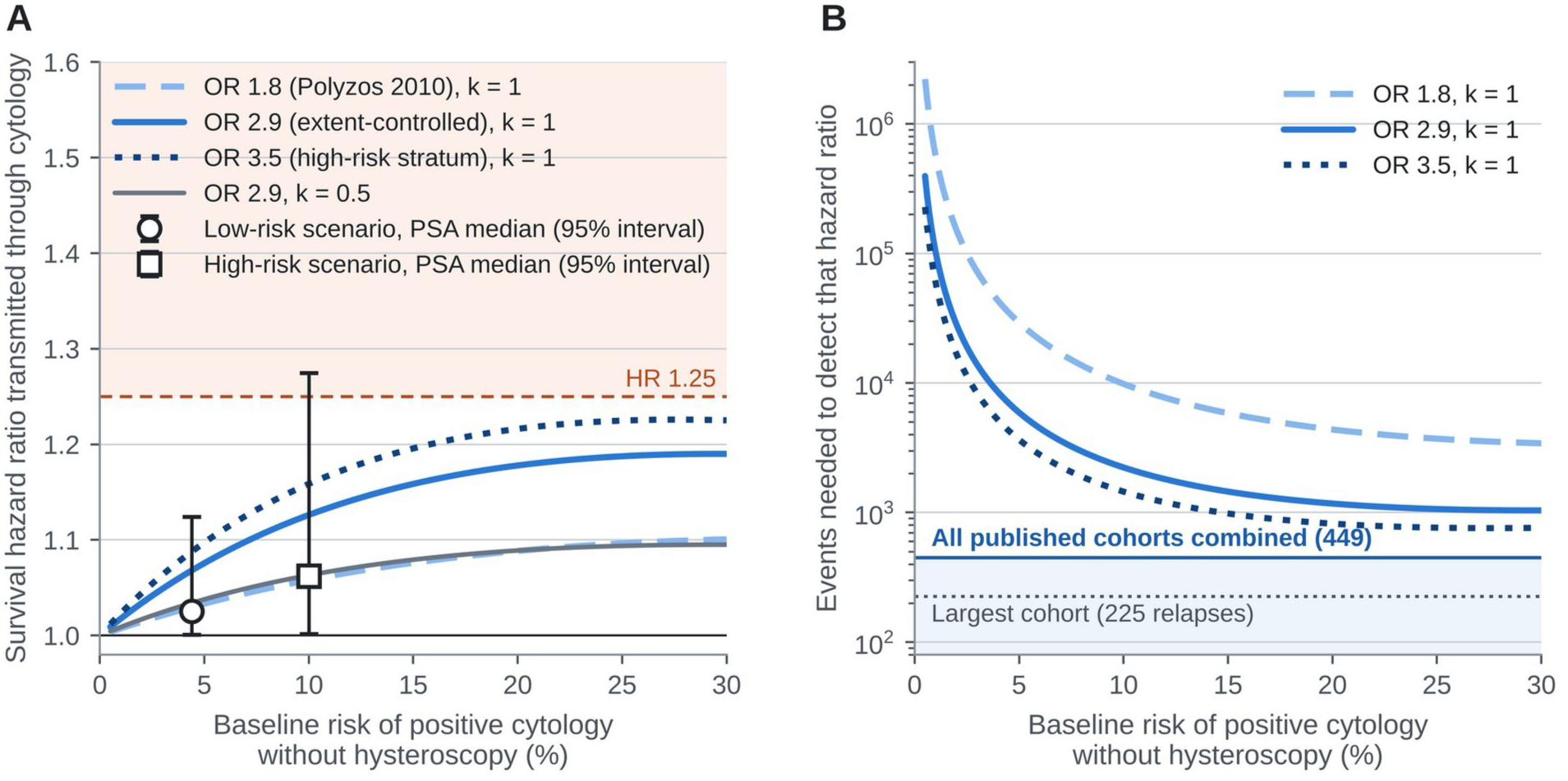
Survival harm that positive cytology could transmit, and the information needed to detect it (Step 6). (A) Hazard ratio transmitted through cytology as a function of the baseline risk of positive cytology, for three ORs and two values of k, the fraction of hysteroscopy-induced positive cytology that carries the prognostic weight of spontaneous positive cytology (HR 1.98). Markers show medians and 95% intervals from probabilistic sensitivity analysis for the low-risk early-stage and high-risk scenarios; the dashed line marks HR 1.25. (B) Events required to detect the transmitted hazard ratio with 80% power at a two-sided α of 0.05; horizontal lines mark the information in all published cohorts combined and the events in the largest cohort. HR, hazard ratio; OR, odds ratio; PSA, probabilistic sensitivity analysis.

#### Step 7: certainty and absolute effects

Certainty was low for positive cytology and very low for recurrence and survival (Table 6). With disease extent controlled, hysteroscopy would add about 72 positive cytology results per 1,000 women with low-risk early-stage cancer (95% CI 10 to 190 more) and 141 per 1,000 with high-risk histology. The pooled hazard ratio corresponds to 8 more recurrences per 1,000 women at a 5-year risk of 5% (5 fewer to 25 more) and 41 more at 30% (26 fewer to 118 more). Harm transmitted through cytology corresponds to a median of 1.2 additional recurrences per 1,000 women with low-risk early-stage cancer (at most 6.0) and 15 per 1,000 with high-risk histology (up to 65). Because these effects apply only to women who have cancer, expected harm per procedure scales with the pretest probability: about 1.4 additional positive cytology results per 1,000 hysteroscopies for an asymptomatic polyp and 31 when sampling has shown atypical hyperplasia, in which concurrent carcinoma was found in 42.6% [69]. Across pretest probabilities from 1.9% to 42.6%, the expected harm per 1,000 procedures ranged from 1.4 to 31 additional positive cytology results and, if the pooled hazard ratio applied, from 0.2 to 3.5 additional recurrences (upper limits 0.5 and 11) (Table 5; S6 Table). The pretest probability therefore changes the balance by a factor of more than 20, which is why the measures in Step 8 are stratified by it.

#### Step 8: recommendations and proportionate precaution

No guideline specifies a distension pressure or medium for women in whom cancer is likely, and none grades the evidence on dissemination (Table 7). The European guideline recommends that intraperitoneal tumor spillage be avoided (level III, grade A) but does not address hysteroscopy or peritoneal cytology [56]. Other oncology guidelines endorse hysteroscopy for diagnosis or infer an unaffected prognosis from nonsignificance without grading [57–59], and hysteroscopy statements favor visually directed sampling without addressing dissemination [20,21]. The staging system of the International Federation of Gynecology and Obstetrics excludes cytology [90,91], whereas the National Comprehensive Cancer Network advises that it be obtained because it is an adverse risk factor [60].

**Table 7.** How current guidelines address hysteroscopy when endometrial cancer is suspected (Step 8)

| Guideline | Role assigned to hysteroscopy | Dissemination or cytology | Pressure or medium | Opportunity identified by the audit |
|---|---|---|---|---|
| ESGO–ESTRO–ESP 2025 [56] | Not addressed | Cytology not addressed; any intraperitoneal tumor spillage should be avoided (III, A) | Not addressed | Extend the spillage principle to diagnostic distension |
| ESMO 2022 [57] | May help to obtain a representative biopsy or remove the lesion (ungraded); biopsy or curettage acceptable (IV, A) | Not addressed | Not addressed | Address the harms of diagnostic procedures |
| German S3, version 4.1, 2026 [58] | Hysteroscopy with fractional curettage is the diagnostic gold standard (statement, evidence level 3) | No statement | No statement | Specify sequence and pressure when cancer is likely |
| BGCS 2022 [59] | If outpatient biopsy is not feasible (B); first-line outpatient hysteroscopy may be considered (D) | Slightly increased positive cytology; prognosis considered unaffected (ungraded) | Not addressed | State the margin of harm excluded, not only the absence of significance |
| AAGL–ESGE–GCH 2026 [20] | Visually directed biopsy preferred when feasible (strong; low to moderate certainty) | Not addressed | Not addressed | Qualify for the scenario of likely cancer |
| FIGO–GCH 2025 [21] | Gradual replacement of blind procedures where feasible | Not addressed | Not addressed | Include oncologic outcomes |
| FIGO staging 2009 and 2023 [90,91] | Not applicable | Cytology excluded from stage assignment | Not applicable | Keep recording cytology for its prognostic value |

Grades as given by each guideline. AAGL = American Association of Gynecologic Laparoscopists; BGCS = British Gynaecological Cancer Society; ESGE = European Society for Gynaecological Endoscopy; ESGO = European Society of Gynaecological Oncology; ESMO = European Society for Medical Oncology; ESP = European Society of Pathology; ESTRO = European Society for Radiotherapy and Oncology; FIGO = International Federation of Gynecology and Obstetrics; GCH = Global Community on Hysteroscopy.

The knowledge condition for precaution was met: there is direct human evidence of transtubal passage of viable tumor cells [25] and a controlled association with an intermediate of known prognostic weight [28]. The harm condition was also met, because recurrence of endometrial cancer is serious and often incurable. Three candidate measures were compared (Fig 9). Abandoning diagnostic hysteroscopy was inconsistent, because it would forgo the detection of focal lesions that blind sampling misses [92,93] in a population most of whom do not have cancer. Using the lowest effective pressure and recording medium and pressure in every procedure was consistent and nearly costless [94]. Performing curettage before hysteroscopy, under the same anesthetic, when cancer is likely and preoperative information is incomplete was consistent and efficient: tissue that looks like tumor makes distension unnecessary, hysteroscopy follows at low pressure when it does not, and the measure is confined to this scenario because curettage carries its own risks (intraoperative complications in 1.9% of 5,359 procedures) [95]. These measures are provisional and proportionate to the present level of uncertainty, and they are tied to the same rule that produced them. They would be withdrawn if a registry-based or randomized comparison established safety under Step 5 at the margin of 1.25, or if induced positive cytology were shown to be prognostically inert; they would be strengthened, for example by avoiding diagnostic distension whenever cancer is likely, if Step 5 established harm. Table 8 summarizes the audit.

**Fig 9.**
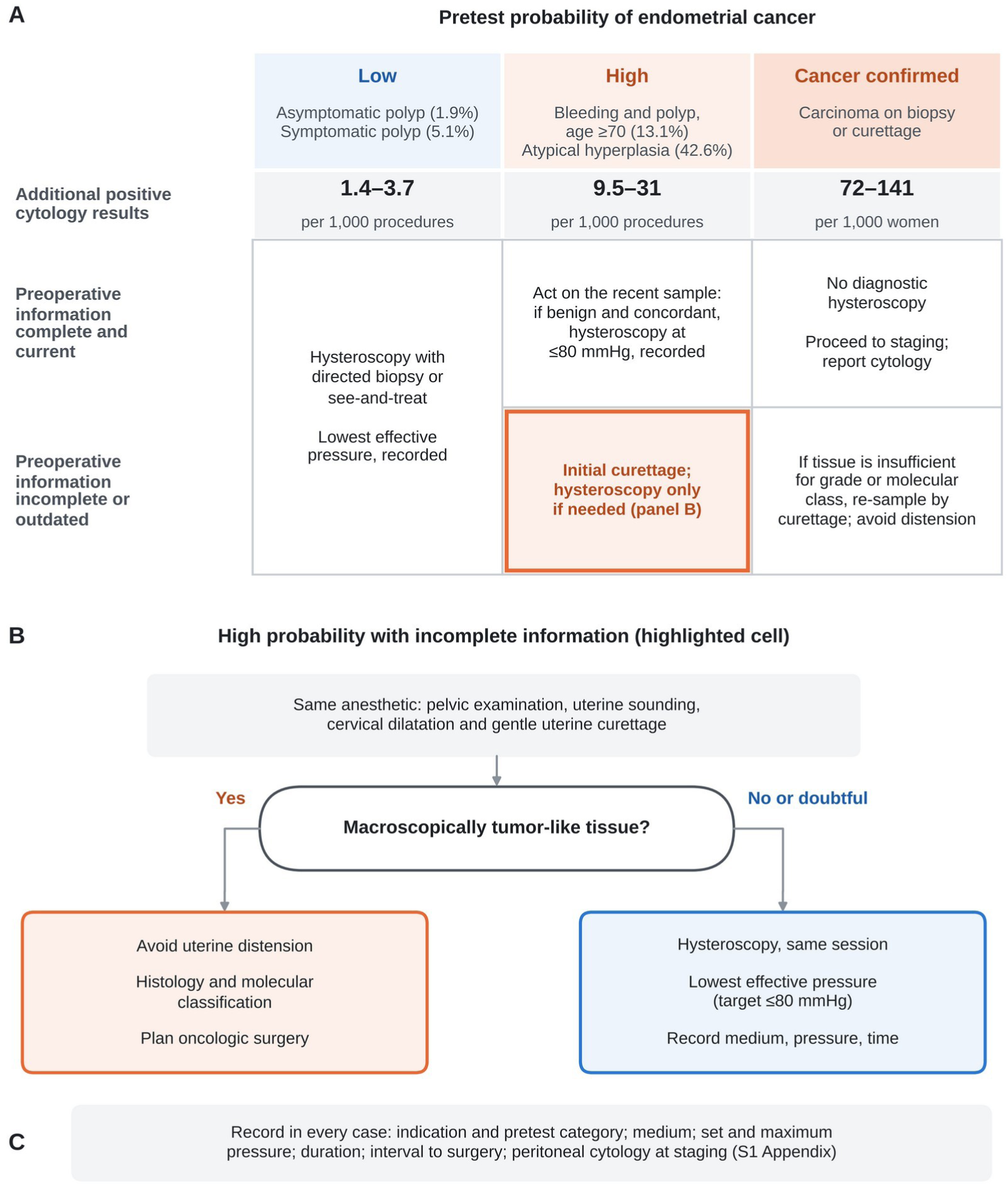
Proportionate precaution by pretest probability of cancer and completeness of preoperative information (Step 8). (A) Each cell gives the proposed action. The Step 5 verdict on recurrence, inconclusive, is the same in every column; the actions differ because the expected harm differs. The examples under each column give the reported probability of cancer in that scenario [23,68,69]; the row beneath gives the expected number of additional positive cytology results per 1,000 procedures, calculated from the extent-controlled odds ratio (S6 Table; for confirmed cancer, per 1,000 women with low-risk or high-risk disease, Table 6). The highlighted cell, high probability with incomplete information, favors initial curettage with conditional hysteroscopy. (B) Same-anesthetic sequence in the highlighted cell: if the curetted tissue looks like tumor, distension is avoided and histologic and molecular classification guide surgery; if not, hysteroscopy follows at the lowest effective pressure. The pressure target of 80 mmHg or less is expert opinion. (C) Items to record in every case (record form in S1 Appendix). The measures are provisional: they are proportionate to current uncertainty and would be withdrawn or strengthened if Step 5 established safety or harm.

**Table 8.** Audit report for the claim that diagnostic hysteroscopy is oncologically safe in endometrial cancer.

| Step | Finding | Verdict or implication |
|---|---|---|
| 1. Claim and margin | Diagnostic hysteroscopy before surgery does not increase recurrence; margin HR 1.25; intermediate, positive peritoneal cytology | Claim testable |
| 2. Provenance | 17 primary estimates: 12 copied, 3 derived, 2 reconstructed; discrepancies in secondary tabulations, including a randomized trial and a unit error | Analyses based on primary reports only |
| 3. Direction of confounding | Controlled OR 2.86 vs crude 0.90; increase after control in 3 of 3 studies; E-value 1.81 to mask HR 1.25 | Crude comparisons biased toward safety; not accepted as evidence of safety |
| 4. Effect scale | 5 cohorts with hazard ratios; published syntheses pooled cumulative proportions | Published risk ratios (0.80, 0.88) not evaluable as evidence of absence |
| 5. Evidence of absence | HR 1.17 (0.90–1.52); 449 event-equivalents (71%); z<sub>harm</sub> 1.65, z<sub>safe</sub> 0.71 vs 2.32; audit interval 0.94–1.46; P(HR > 1) 91%, P(HR > 1.25) 26% | Inconclusive at margins from 1.10 to 1.40; only an increase of 50% or more excluded |
| 6. Detectability | Transmitted HR at most 1.07 (low risk) and 1.16 (high risk); 7,300–50,000 and about 1,400 events needed | Absence of a survival signal uninformative in low-risk disease |
| 7. Certainty and absolute effects | Cytology: low certainty, 72 more per 1,000; recurrence: very low certainty, 8 more (5% risk) to 41 more (30% risk) per 1,000; per 1,000 procedures, 1.4 to 31 more positive cytology results, depending on the pretest probability | Harm not excluded; largest in high-risk histology |
| 8. Recommendations and precaution | No guideline addresses pressure or grades dissemination; knowledge and harm conditions met | Documented low-pressure hysteroscopy in all (1.4 to 3.7 more positive cytology results per 1,000 procedures when cancer is unlikely); curettage first when cancer is likely (9.5 to 31 per 1,000 procedures) and information incomplete; no diagnostic hysteroscopy once cancer is confirmed (72 to 141 per 1,000 women) |
| Conditions for revision | Same Step 5 rule applied to new registry-based or randomized evidence | Withdraw if safety is established or induced cytology is shown to be inert; strengthen (avoid distension when cancer is likely) if harm is established |

HR = hazard ratio; OR = odds ratio. Full results in the text, Table 5 and S4 Table; a blank worksheet and a one-page committee summary for other claims are provided in S1 File.

## Discussion

### Principal findings

This study proposes an eight-step framework for auditing claims of safety and shows three things. First, the rule most often used in practice, inferring safety from nonsignificance, gives false reassurance in most scenarios in which a clinically important harm exists and evidence is limited (34% to 91% of analyses in the scenarios examined), whereas a symmetric, information-adjusted rule keeps this error at or below its nominal 2.5% for a single analysis and still establishes safety when information is adequate (80% of analyses at the required information when there is no harm). Second, applied with the rule locked in advance to the evidence available before the trial, the framework would have classified minimally invasive radical hysterectomy as unproven rather than safe, and the reassurance offered by the pre-trial meta-analysis would have failed at four separate steps. Third, applied to diagnostic hysteroscopy, the framework shows that the evidence neither establishes nor excludes oncologic harm: an association with positive cytology emerges once disease extent is controlled, time-to-event data lean toward harm without crossing either boundary, only a large increase in recurrence is excluded, and harm transmitted through cytology lies beyond the detection limit of any feasible cohort in low-risk disease. Taken together, the findings imply that a claim of safety should be accepted only when it states the margin of harm it excludes and rests on enough verified information to exclude it. In practice, the central check takes minutes: with the two limits of a published 95% confidence interval, Table 2 gives the Step 5 verdict and what it means for decisions.

### Comparison with existing methods

Each component draws on established methods; the contribution lies in their integration around a single question and in elements that existing tools do not provide. Trial sequential analysis relates accrued to required information [12], and the eight-step procedure of Jakobsen and colleagues extends this to statistical and clinical significance in meta-analyses of randomized trials [13]; neither targets claims of absence of harm built on observational data. The audit interval of Step 5 is not new as a statistical object, because adjusted and repeated confidence intervals serve the same purpose in sequential analysis [12,42]; what is new is its use, with a margin set in advance, as a symmetric test of claims of safety and harm.

GRADE, ROBINS-I and equivalence methods each supply part of what a claim of safety requires. GRADE rates certainty and treats as imprecise an interval that includes appreciable harm or rests on less than the optimal information size [9,10], but it returns a certainty rating rather than a verdict on the claim and leaves to the panel how much uncertainty justifies precaution. ROBINS-I judges each nonrandomized study and can record the predicted direction of bias [11], but it neither verifies the numbers nor tests that prediction against crude and controlled estimates. Equivalence tests [14] and interval classifications such as second-generation P values [96] give a verdict against a prespecified margin, and at full information Step 5 reduces to a one-sided equivalence test; but their critical value ignores how much of the required information has accrued, so a margin check applied repeatedly to accumulating evidence declared safety at some update in 9.7% of simulated bodies of evidence with harm at the margin. The framework adds what none of these provides on its own: one verdict, symmetric for harm and safety, against a margin set in advance and adjusted for the information accrued; verification of each number against its source (Step 2), a test of the predicted direction of confounding (Step 3), the time-to-event scale for time-dependent outcomes (Step 4) and an assessment of whether the harm was detectable at all (Step 6); and a precaution tied to the verdict, with the evidence that would withdraw or strengthen it stated beforehand (Step 8).

A reader might ask what the framework adds to applying GRADE, trial sequential analysis and an equivalence test separately to the same evidence. Together these supply most of the statistical ingredients of Step 5, but not a verdict, and applied separately they can disagree without a rule to reconcile them. For the pre-trial meta-analysis of minimally invasive radical hysterectomy, an equivalence test at a margin of 1.25 accepted safety, trial sequential analysis would have shown that only 27% of the required information had accrued, and GRADE would have returned low or very low certainty. Trial sequential analysis comes closest, because Step 5 uses its boundary for a single analysis. But trial sequential analysis is built to test benefit or harm, and its futility boundaries, the usual way to conclude that an effect of a given size is absent, are calibrated by β-spending, so they accept a probability of false reassurance at the anticipated effect of up to the chosen β, commonly 10% or 20% [12,97]; Step 5 holds it at or below 2.5% for a single analysis, the standard it also applies to a claim of harm. None of the three verifies the numbers against the primary reports, tests the predicted direction of confounding, rejects pooled cumulative proportions for time-dependent outcomes or asks whether the harm could have been detected; in the retrospective case each of Steps 2 to 4 found a problem that these tools do not test (Table 3). The framework therefore does not replace them. It fixes the order in which they are used, adds the checks they omit, and reduces their outputs to one prespecified verdict that is kept apart from the decision about precaution.

The pre-trial evidence on minimally invasive radical hysterectomy shows what this adds in practice (Table 9). Read for statistical significance, the meta-analysis and the matched cohort showed no difference, and their authors concluded that the approach was safe [46,47]; a margin check would have accepted the meta-analysis, because its upper limit of 1.11 lay below 1.25; by our reconstruction, not a formal application of either tool, GRADE would have rated the evidence at low or very low certainty without a verdict on the claim [9,10]; and ROBINS-I would have rated each study without testing the direction of its bias [11]. Step 5 classified the same evidence as inconclusive: its audit interval, 0.46–1.46, left increases of up to 46% compatible with the data, and Steps 2 to 4 flagged problems in the numbers, the direction of bias and the effect scale that the other approaches do not test. The trial later reported a hazard ratio for recurrence or death of 3.74 [48]. The framework could not have anticipated that result, and one case selected by us cannot show how often it reads evidence correctly; what it changed was the reading of the pre-trial evidence, from “safe” to “not shown to be safe”, with the information needed to settle the question stated.

**Table 9.** How each approach would have read the evidence available before the randomized trial of minimally invasive radical hysterectomy; the readings attributed to GRADE, ROBINS-I and trial sequential analysis are the authors’ reconstruction, not formal applications of these tools.

| **Approach** | **Reading of the pre-trial evidence** | **What it leaves open, or what the framework adds** |
|---|---|---|
| Statistical significance | No significant difference in the meta-analysis (OR 0.82, 0.61–1.11) or the matched cohort (HR 1.28, 0.62–2.64); the authors concluded that the approach was safe [46,47] | Absence of evidence read as evidence of absence; no margin of harm stated |
| Margin check or one-sided equivalence test (margin 1.25) | Safety accepted for the meta-analysis: upper limit 1.11, below 1.25 (z<sub>safe</sub> 2.76 against 1.96) | Ignores that only 27% of the required information had accrued |
| GRADE (authors' reconstruction) | Starts at low certainty; could be rated down to very low for imprecision [9,10] | A certainty rating, not a verdict on the claim; the conventional interval appears to exclude increases above 11% |
| ROBINS-I (authors' reconstruction) | Risk of bias judged study by study; main concern, confounding by tumor size [11] | Does not test the predicted direction of bias or judge the pooled claim |
| Trial sequential analysis (authors' reconstruction) | 27% of the required information (171 of 631 event-equivalents); boundary for harm not crossed | Absence of harm is concluded only at futility boundaries calibrated by β (often 20%), not α; numbers, direction of bias and effect scale not examined |
| **This framework (Steps 2 to 5)** | **Inconclusive: not shown to be safe; 171 of the 631 event-equivalents required; audit interval 0.46–1.46** | Increases of up to 46% remain compatible; inconsistent effect labels, confounding toward safety and pooled cumulative proportions flagged; the information needed to settle the question is stated |

The randomized trial later reported a hazard ratio for recurrence or death of 3.74 (1.63–8.58) [48], and subsequent syntheses established harm under Step 5 (Table 3) [50,51]. The readings attributed to GRADE, ROBINS-I and trial sequential analysis are the authors' reconstruction of how these tools would apply; none was applied formally. GRADE = Grading of Recommendations Assessment, Development and Evaluation; HR = hazard ratio; OR = odds ratio; ROBINS-I = Risk Of Bias In Non-randomised Studies of Interventions; β = type II error.

### Absence of evidence, evidence of absence and precaution

The distinction between absence of evidence and evidence of absence is long established [1], and equivalence testing gives it a formal basis [14]. The framework makes the distinction operational for bodies of observational evidence that are updated over time, and separates it from the decision that follows. Step 5 is deliberately symmetric and presumes neither harm nor safety; precaution enters only at Step 8, only when its conditions are met, and only through measures shown to be proportionate. An inconclusive verdict therefore does not dictate a measure; it leads to Steps 7 and 8, where the harm at stake if the effect is real, expressed per 1,000 patients or procedures, determines which measure, if any, is proportionate (Table 2; Fig 9). This separation answers the principal criticism of precautionary reasoning, that it can justify any restriction [17]: the evidential verdict does not depend on who applies it, and the recommendation must show that it is not ruled out by the risks it creates [18,19]. The retrospective case illustrates the division of labor. A single truncated trial provided a strong signal that met the knowledge condition for precaution but not, by itself, an established effect size, consistent with the tendency of trials stopped early to overestimate effects [98] and with the smaller hazard ratios reported by later syntheses [50,51].

### Implications for clinical practice

Two outputs of the audit should be kept apart. The verdict is a statement about the evidence: for diagnostic hysteroscopy in endometrial cancer, oncologic harm is neither shown nor excluded (Step 5). The precautions are decisions about what to do while that question remains open (Step 8): they are justified because the possible harm is plausible and serious, they are scaled to the harm expected if the effect is real, and they do not imply that harm has been shown. A reader may therefore disagree with a particular precaution without disputing the verdict, and the precautions are withdrawn or strengthened by the same rule that produced the verdict as evidence accumulates. Table 10 makes the division explicit: for the verdict and for each precaution it states whether the output was computed by the locked rule or judged, what it rests on, how it behaved when its key assumptions were varied, and what would change it.

**Table 10.** Verdict and recommendations for diagnostic hysteroscopy: what each rests on and how sensitive it is to the assumptions.

| Output (step) | Computed or judged; what it rests on | Behavior when key assumptions are varied | What would change it |
|---|---|---|---|
| **Verdict: inconclusive (Step 5)** | Computed by the locked rule from the pooled hazard ratio, 1.17 (0.90–1.52), and the margin; no other input | Inconclusive at every margin from 1.10 to 1.40, with recursively calibrated boundaries, and in each sensitivity set (Table 5; Fig 5); only an increase of 50% or more is excluded, narrowly | A registry-based or randomized comparison that crosses a Step 5 boundary |
| **Lowest effective pressure, with medium and pressure recorded, in every diagnostic hysteroscopy (Step 8)** | Judged: knowledge and harm conditions met; nearly costless [94] | Independent of the pretest probability and the baseline risks; if induced cytology were prognostically inert (k = 0), it would rest only on the direct time-to-event estimate | Withdrawn if safety is established at the margin or induced cytology is shown to be inert |
| **Target of 80 mmHg or less (Step 8)** | Expert opinion; indirect support from published subgroups, OR 1.18 below 80 mmHg and 2.18 above [27] | Not derived from Step 5; the subgroup contrast is indirect and imprecise | Comparisons of cytology or recurrence by recorded pressure |
| **Curettage first when cancer is likely and preoperative information is incomplete (Step 8)** | Judged: 9.5 to 31 additional positive cytology results per 1,000 procedures, against 1.4 to 6.5 when cancer is less likely; curettage has its own risks (complications in 1.9%) [95] | Expected harm changes more than 20-fold with the pretest probability (Table 5); placing the threshold between 9.0% and 13.1% is a judgment; with k = 0 there is no survival harm through cytology | As above; strengthened (avoid distension whenever cancer is likely) if harm is established |
| **No diagnostic hysteroscopy once cancer is confirmed (Step 8)** | Judged: 72 to 141 additional positive cytology results per 1,000 women with cancer; little added to staging | Rests on the extent-controlled OR, 2.86 (1.24–6.62), for cytology and on k for any survival effect; independent of the pretest probability | As above |

Only the verdict is produced by the locked rule; every precaution also depends on judgments about baseline risks, pretest probabilities, the prognostic weight k of induced positive cytology and the risks of each measure, and a reader may accept the verdict while weighing these differently. Sensitivity results in Table 5 and Fig 8. k = fraction of hysteroscopy-induced positive cytology carrying the prognostic weight of spontaneous positive cytology; OR = odds ratio.

For practice, the audit supports a stratified approach based on the pretest probability of cancer and the completeness of preoperative information, rather than a uniform rule (Fig 9): the Step 5 verdict is the same in every setting, but the expected harm is not, and each measure is scaled to it. When the probability of cancer is low, as for an asymptomatic or symptomatic polyp (1.9% and 5.1%) [68], hysteroscopy with directed biopsy or see-and-treat can proceed as usual, at the lowest pressure that gives an adequate view and with medium and pressure recorded; the expected harm is small per procedure (1.4 to 3.7 additional positive cytology results per 1,000 procedures). When cancer is likely, as in women aged 70 years or older with bleeding and a polyp (13.1%) or after sampling has shown atypical hyperplasia (42.6%) [23,69], the expected harm is larger (9.5 to 31 additional positive cytology results per 1,000 procedures), and the decision depends on preoperative information. If a recent sample is benign and concordant with imaging, hysteroscopy can follow at 80 mmHg or less and be recorded; if information is incomplete or outdated, curettage under the same anesthetic comes first, distension is avoided when the tissue looks like tumor, and hysteroscopy follows at low pressure only when it does not. When cancer has already been confirmed, the excess applies to every woman examined (72 more positive cytology results per 1,000 women with low-risk early-stage disease and 141 with high-risk histology), and diagnostic hysteroscopy adds little to staging, which relies on imaging and surgical pathology; if the sample is insufficient for grade or molecular classification, re-sampling by curettage avoids distension. The pressure target of 80 mmHg is expert opinion, and these measures are conditional: they are proportionate to current uncertainty and would change, in either direction, if new evidence crossed a Step 5 boundary.

What changes in practice, and what does not, can therefore be stated precisely. The audit gives no reason to abandon hysteroscopy, a measure rejected at Step 8 because it would forgo the detection of focal lesions that blind sampling misses, and it does not alter the indications for hysteroscopy, staging or treatment. What changes is how the procedure is performed and recorded when cancer is possible, the order of sampling when cancer is likely and preoperative information is incomplete, and how its oncologic safety is described: as not established, with the margin that was excluded and the information on which that statement rests. A hospital protocol could state: “When endometrial cancer is likely, for example after atypical hyperplasia on sampling or with bleeding and a polyp at age 70 years or older, and preoperative information is incomplete or outdated, perform curettage first under the same anesthetic; avoid distension if the tissue looks like tumor, and otherwise perform hysteroscopy at the lowest pressure that gives an adequate view (80 mmHg or less; expert opinion). In every diagnostic hysteroscopy, record medium, pressure, duration and the interval to surgery. This protocol will be reviewed if new evidence establishes safety or harm.” Each element would be withdrawn or strengthened under the conditions stated in Table 8, and S1 File provides a one-page committee summary that brings together the verdict, what changes, what does not and the conditions for revision.

Related findings support this approach without settling the question. The mechanism invoked, intrauterine pressure driving tumor cells through the tubes, also underlies concern about uterine manipulators, for which a meta-analysis estimated a hazard ratio for disease-free survival of 1.18 (1.01–1.38) [4], close to the pooled estimate for hysteroscopy. Neither the duration of hysteroscopy nor the interval to surgery has been associated with positive cytology [74,99], no transfer of endometrial cells was observed with carbon dioxide at a controlled 80 mmHg [55], and hysteroscopic tissue removal was not associated with greater dissemination in one cohort [100]. For counseling, a plain statement is supported and can be used as written: “Hysteroscopy is accurate and usually well tolerated. When cancer is present, it may increase the chance of finding cancer cells in abdominal washings, and current studies cannot exclude a small increase in the risk of recurrence, which would matter most for aggressive tumors.”

### Generalizability, independent use and adoption

The questions in Table 1 can be asked of any claim that an intervention is safe, without software: what margin of harm was excluded; whether the numbers match the primary reports; which way the selection of patients would bias the comparison; whether time-dependent outcomes were analyzed over time; whether there was enough information to detect the margin; and whether the harm could have been detected at all. Because the rule’s error rates depend only on the true effect, the margin and the information available, not on the clinical question (Fig 2; S1 Appendix), its protection against false reassurance carries over to other claims of safety based on ratio estimates; what must be set anew for each claim is the margin, by clinical judgment. The other steps transfer to different degrees (S1 Appendix, section H). Steps 1 to 5 and 7 apply to any claim of safety that rests on comparative estimates; Step 6 applies when the harm is thought to act through an intermediate whose prognostic weight is known, and is otherwise recorded as not applicable; and Step 8 depends on the clinical setting, the alternatives available and the values of those affected, so the precautions proposed for hysteroscopy are a result of this audit, not a template for other interventions. The steps can also be used separately: Step 5 needs only a ratio estimate, its confidence interval and a margin, and Step 2 can be applied by anyone extracting data for a synthesis. The same procedure could be applied to other claims of oncologic safety, such as those concerning uterine manipulators [4] or the de-escalation of surgical staging, and to claims about drugs and devices that rest on observational comparisons. For the 2022 meta-analysis of uterine manipulators cited in the Introduction [3], Table 2 gives the verdict in a few seconds: the ratio of the limits, 1.74 ÷ 0.71 = 2.45, falls in the row for 2.50, and neither threshold is met, so the evidence was inconclusive, with about 76 event-equivalents, 12% of the information required; its risk ratio also pooled recurrence proportions, which Step 4 does not accept as evidence of absence. We do not presume the results of such audits: the framework is designed to report safety when the evidence supports it as readily as it reports uncertainty or harm.

Because the framework was developed on the hysteroscopy evidence, the risk that it was fitted to that case needs explicit attention. Three features limit this risk. The part that decides the verdict, the Step 5 rule, has no parameter estimated from data: its constants, the GRADE threshold of 1.25, 80% power, a two-sided α of 0.05 and an O’Brien–Fleming-type boundary, are conventional, and the rule was locked before it was tested. Its error rates were derived analytically and by simulation for any claim based on a ratio estimate, not estimated from the hysteroscopy data (Fig 2). And the steps that involve judgment produce explicit outputs, a provenance grade for every number, a stated direction of bias and the conditions attached to each measure, that others can check against the primary reports. The computations can also be reproduced without the developers: the version that needs no software gave the same verdict as the full calculation for each of the 9 confidence intervals audited here, the complete analysis was reproduced on a separate computer by an author who had not conducted it (AR), and the calculator reproduces the values reported here in its built-in self-check. What these features cannot exclude is that the steps were shaped by the problems this particular body of evidence presented, and two applications cannot show how often the framework reads evidence correctly; only applications by others, to claims they select, can show that.

Independent applications should therefore be designed so that their results can count against the framework. Each should record the claim and the margin, with its basis, before the evidence is examined and not change the margin afterwards; have two people extract and grade every number against the primary reports; apply the locked rule unchanged, with recursively calibrated boundaries when the same evidence has been audited before; record which entries were computed and which were judged; and deposit the completed worksheet with the report (S1 File). Three studies would then test what the present evidence cannot. First, agreement: auditors who did not develop the framework would apply it independently to the same published claims, and agreement on step-level findings and verdicts would be measured. Second, predictive performance: people other than the developers would select, by predefined criteria, claims of safety based on observational evidence that were later tested in randomized trials or large registries, and apply the locked rule to the evidence available before each test, ideally before its results are known and otherwise blind to them; this would remove the selection by the developers that limits the present retrospective test. Third, effect on decisions: after usability testing, clinicians or guideline methodologists would be randomly assigned to decide on clinical vignettes with or without the framework’s outputs, and their decisions and wording compared. The framework should be revised or restricted if auditors applying the same protocol often reach different verdicts, if claims it classifies as safety established are later shown to carry harm beyond the margin more often than its nominal error rate of 2.5% allows, or if its outputs do not improve decisions or the wording of claims.

The least demanding form of adoption does not depend on these studies, because it changes only how claims are stated. Journals, reviewers and guideline panels can ask of any claim that an intervention is safe for the margin of harm it excludes, the estimate with its 95% confidence interval and the Step 5 verdict read from Table 2, and wording that matches the verdict, so that a nonsignificant difference is not reported as evidence of safety; S1 File gives wording for instructions to authors and review templates, and a full audit can be reserved for claims that would change practice. Table 11 sets out a path from a single published interval to a full audit and to the evaluation of the framework, with what each user needs and where it is found; each level can be used on its own and leads to the next.

**Table 11.**
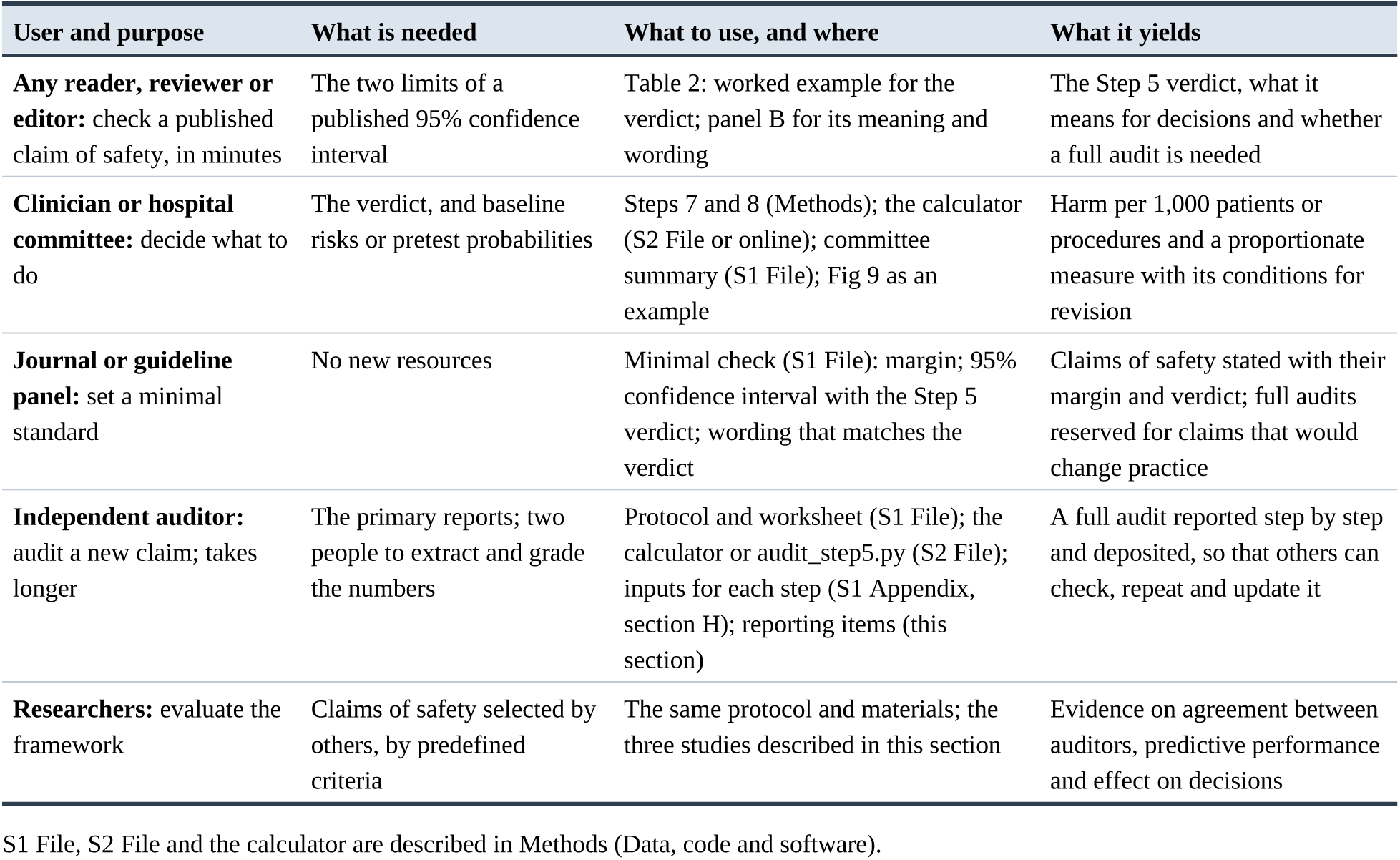
A path for using and adopting the framework: who, what is needed, what to use and what it yields.

When the framework is applied to another claim, the report should allow others to repeat the audit. We suggest reporting the items that the worksheet in S1 File records for each step: the claim as stated and where it is made; the margin, with its basis and whether it was set before the evidence was examined; the provenance grade of every number and any discrepancy found; the predicted direction of confounding and the design feature used to test it; the effect measure of each estimate; for Step 5, the type and level of each interval, how its standard error was obtained, the information available and required, the boundary, the audit interval and the verdict, with any earlier audits of the same evidence; the assumptions behind the detectability bound; the certainty ratings and the baseline risks and pretest probabilities behind the absolute effects; and the measures adopted and rejected, with the evidence that would withdraw or strengthen each. The list is our proposal, not a reporting guideline developed by consensus.

### Implications for guidelines and research

The audit suggests that guidance on this question could be made more explicit at little cost. Guidelines could state the margin against which safety is judged, grade evidence on harm separately from evidence on diagnostic accuracy, and extend the principle of avoiding tumor spillage, already endorsed for surgery [56], to diagnostic distension, for example by recommending the lowest distension pressure that provides adequate visualization when endometrial cancer is suspected or confirmed, with medium, pressure, duration and the interval to surgery recorded in the operative note. Removing cytology from surgical staging did not remove its prognostic weight [28,101,102], so recording it at staging remains informative.

GRADE separates the certainty of evidence from the strength of a recommendation [103], and strong recommendations based on low-certainty evidence call for explicit justification [104]; the issue here is not the strength of current recommendations but explicitness about what has and has not been shown. Claims of safety can be worded to state what was excluded and with how much information (Table 2, panel B), and recommendations that rest on inconclusive evidence can state in advance what would change them (Table 8), so that revision follows from applying the same rule to new evidence rather than from a new debate. A randomized noninferiority trial with a margin of 1.20 would need about 944 events, roughly 9,400 women at a 10% recurrence risk, which is unlikely to be feasible. Registry-based emulation of a target trial that records pressure, medium, duration and interval to surgery is the realistic path [105], and reporting hazard ratios with confidence intervals rather than proportions would allow evidence to accumulate [34]; for syntheses that are updated repeatedly, the recursively calibrated boundaries keep the overall error at its nominal level.

### Strengths and limitations

The framework is explicit, uses only published data and is fully reproducible from the files provided: every number carries a provenance grade, every step has a stated output, and one command regenerates every result. Its decision rule contains no parameter estimated from data, was locked before the retrospective test, and has operating characteristics derived analytically, independently of any data set. The same rule can report safety, harm or uncertainty, and the outputs, a provenance log, a verdict and a revisable recommendation, can be checked and repeated by others, with or without software.

The main limitation is that the framework was developed on the case to which it is applied, which risks fitting the method to the conclusion. We mitigated this by locking the Step 5 rule before the retrospective test, by using only conventional constants in it, by evaluating its operating characteristics independently of any data set, and by testing it on a second case; because no protocol was registered, independent application to other claims is the necessary next step, and S1 File provides a protocol for it. For the same reason, the hysteroscopy application illustrates the framework but provides no evidence that the rule performs well; that evidence comes from its operating characteristics and, within the limits of a single case, from the retrospective test, which used pre-trial evidence chosen by us and cannot estimate sensitivity or specificity. The usability of the materials, the agreement between auditors who apply the framework independently to the same claim, its effect on decisions and its performance on claims selected by others have not yet been evaluated; the studies needed are described above (Generalizability, independent use and adoption) and in S1 Appendix, section G. External validation is therefore absent: the framework has not been applied prospectively, by auditors independent of its developers, or to claims selected by others, so it is not known how reproducible the steps that involve judgment (Steps 2, 3, 6, 7 and 8) are between auditors, nor how often the complete procedure reads evidence correctly. The operating characteristics reported here apply to the Step 5 rule, not to the eight-step procedure as a whole. Until these evaluations are available, the framework should be regarded as a provisional proposal.

The verdict itself has limitations. The locked boundary does not adjust later analyses for earlier ones, which explains the 3.5% rather than 2.5% error with 10 updates; recursively calibrated boundaries restore the nominal level at a modest cost in power and changed no verdict here. The rule is conservative for single small studies in both directions, and the margin of 1.25 is a convention, although the verdict was unchanged for margins from 1.10 to 1.40. Step 5 is specified for ratio measures whose confidence intervals are symmetric on the log scale; applying the same structure to absolute differences, with the margin and the information defined on that scale, was not evaluated.

The evidence on hysteroscopy has its own limitations. The search was structured, updated and reported according to PRISMA 2020, but it did not query every bibliographic database directly, the application was not registered, and ROBINS-I was not applied study by study. The grouping of cytology studies by control of disease extent was defined during the analysis, and the difference between groups (P = 0.04) rests on few studies, so its interpretation relies mainly on the consistent within-study pattern. Several estimates were derived or reconstructed, although provenance grading makes each step auditable. The mediation bound assumes proportional hazards and borrows the prognostic weight of cytology from registry data. No study reported outcomes by molecular class or recorded pressure and medium consistently.

The precautionary recommendations rest on more judgment than the verdict. Absolute effects depend on assumed baseline risks and pretest probabilities, certainty ratings involve judgment, the knowledge and harm conditions and the comparison of candidate measures are judgments made by the authors, and the pressure target of 80 mmHg is expert opinion. These judgments determine which measures are proportionate but do not change the Step 5 verdict; the calculator computes Step 5 and the absolute effects of Step 7 but leaves these judgments to its user. Table 10 shows which recommendation depends on which assumption: a reader who places the threshold of pretest probability elsewhere, prefers another pressure target or assumes a different value of k may reach different recommendations from the same verdict, which is why the recommendations are reported separately from it.

### Conclusions

Claims of safety should be judged against a stated margin, on verified numbers, on the appropriate effect scale and with adequate information. The framework presented here makes these requirements explicit and reproducible. It would not have endorsed the pre-trial claim of safety for minimally invasive radical hysterectomy, and it shows that the evidence on diagnostic hysteroscopy in endometrial cancer is a case of absence of evidence rather than evidence of absence. That verdict describes the evidence; the precautions that follow from it, documented low-pressure hysteroscopy preceded by curettage when cancer is likely and preoperative information is incomplete, are a proportionate response to the uncertainty it leaves, to be revised under the same rule as evidence accumulates. A table reads the Step 5 verdict from the two limits of any reported confidence interval in minutes, and a worksheet, a one-page committee summary and a calculator that runs in any web browser, with the data and code, let others check, update or extend the audit. Whether these outputs improve decisions has not yet been evaluated, and the framework has been applied only to the two cases reported here; until independent and prospective applications are available it should be regarded as provisional, and the protocol and evaluation studies proposed in the Discussion are designed so that others can test it on claims of their own choosing.

## Data Availability

The data set and code that reproduce every result are available in S2 File and archived at Zenodo (https://doi.org/10.5281/zenodo.24059401). The interactive calculator is available at https://evidenceofharm.netlify.app/. This study reanalyzed published aggregate data no new primary data were collected.

## Abbreviations

CI: confidence interval
GRADE: Grading of Recommendations Assessment: Development and Evaluation
HKSJ: Hartung–Knapp–Sidik–Jonkman
HR: hazard ratio
OR: odds ratio
PRISMA: Preferred Reporting Items for Systematic Reviews and Meta-Analyses
ROBINS-I: Risk Of Bias In Non-randomised Studies of Interventions
SE: standard error.

